# The Impact of Menopause on Hypercholesterolemia and Comorbidities: A Population-Based Study

**DOI:** 10.1101/2024.12.29.24319724

**Authors:** Alberto Esteban-Medina, Víctor de la Oliva, Patricia Fernández del Valle, Ana Sánchez, M. Belen Susin, Carlos Loucera, Joaquín Dopazo, Guillermo Antiñolo

**Affiliations:** Andalusian Platform for Computational Medicine, Andalusian Public Foundation Progress and Health-FPS, Seville, Spain; Institute of Biomedicine of Seville, IBiS, University Hospital Virgen del Rocío/CSIC/University of Sevilla, 41013 Sevilla, Spain; Centre for Biomedical Network Research in Rare Diseases (CIBERER), Seville, Spain; Fetal, IVF and Reproduction Simulation Training Centre (FIRST), Seville, Spain; Department of Surgery, University of Seville, Seville, Spain

## Abstract

**Background:** Hormones play a pivotal role in women’s health. Estrogen, for example, not only regulates reproduction but also protects against several chronic conditions, such as heart disease and osteoporosis. As women age, the transition to menopause—a period characterized by a significant drop in estrogen levels—triggers physiological changes that can affect their overall health and quality of life. These changes extend beyond typical menopausal symptoms like hot flashes and vaginal dryness, potentially leading to increased cardiovascular risk, cognitive decline, and bone density loss.

Hypercholesterolemia is a critical risk factor for atherosclerotic cardiovascular disease and is associated with significant comorbidities, particularly in postmenopausal women. Understanding the sex-specific differences in the progression and impact of hypercholesterolemia is essential for improving patient outcomes.

**Objective:** This study explores the implications of hormonal transition, particularly menopause, on women’s health and highlights the potential of real-world data (RWD) to address these gaps. By integrating sex and gender considerations into clinical decision-making. To achieve so, sex-based differences in the prevalence, progression, and comorbidity patterns of hypercholesterolemia are analyzed at region-wide level, using real-world data (RWD) from the Andalusian Population Health Database (BPS).

**Methods:** A retrospective cohort of 557,034 patients diagnosed with hypercholesterolemia between 2016 and 2022 was analyzed. Patients were stratified by sex and age groups (<50 and ≥50 years). A comparative analysis was performed to assess differences in the prevalence of hypercholesterolemia and associated comorbidities.

**Results:** Women were diagnosed later than men, with an average age of 59.1 years compared to 56.0 years for men. Postmenopausal women exhibited a sharp rise in hypercholesterolemia prevalence, surpassing men in older age groups. Women also displayed higher rates of comorbidities, including osteoporosis, anxiety disorders, and hypertension, particularly during the menopausal transition. The analysis revealed a significant influence of estrogen decline on lipid profiles and comorbidities risk in women. Conversely, men had higher hypercholesterolemia prevalence in younger age groups, likely due to lifestyle and genetic factors.

**Conclusions:** This study demonstrates that menopause is a critical period influencing the progression of hypercholesterolemia and associated comorbidities in women. These findings underscore the importance of sex-specific approaches in the prevention and management of hypercholesterolemia. The use of RWD allowed for a comprehensive evaluation of sex and age disparities, providing actionable insights for clinical practice.

## Introduction

Historically, medicine has relied on pattern recognition largely shaped by male-normative perspectives, which often neglect the diversity of human physiology. This bias has hindered a comprehensive understanding of sex- and gender-based differences in health [1]. Recent studies report higher burden of morbidity-driven conditions in females compared to males, pointing to an urgent need for policies to be based on sex-specific and age-specific data [2]. Aging presents distinct challenges for men and women. Women, while generally living longer, experience higher rates of chronic diseases and multimorbidities [3]. Hormonal, biological, and social factors significantly contribute to these health disparities, yet knowledge gaps remain [4]. Fertility is not just a biological process, it is a fundamental aspect of women’s health throughout their lives. It is essential to rediscover, redefine, and understand female fertility in a broader context, particularly in relation to overall women’s health. Fertility is not limited to reproduction, it is an indicator of the body’s overall functioning and female physiology [5]. Therefore, it is reasonable to consider that fertility, normal ovarian function, and consequently, the production of estrogens, the primary female sex hormones, play a central role in women’s physical and psychological well-being. When fertility is lost, the greatest challenge is the decrease in estrogen, which is the “engine” of women’s health. These hormones are essential for the female body, and their role extends far beyond reproduction.

The intersection of hormonal changes and cardiovascular health constitutes an excellent scenario to understand the disparities in disease progression between men and women. The decline in estrogen during menopause significantly impacts lipid metabolism, contributing to adverse changes in cholesterol profiles and an increased risk of hypercholesterolemia [6]. Estrogen’s protective effects on cardiovascular health are well-documented, and its loss during menopause exacerbates vulnerability to conditions like atherosclerotic cardiovascular disease [3]. These hormonal shifts highlight the critical link between fertility loss and the progression of hypercholesterolemia in women, underscoring the importance of sex-specific approaches to its management and treatment.

Understanding the sex-specific differences in its progression, associated comorbidities, and management is essential for effective treatment strategies. From childhood, females with familial hypercholesterolemia (FH) exhibit higher total cholesterol and LDL-C levels compared to males. By age 30, women with FH have a greater cumulative LDL-C burden [7]. Despite this, women are often diagnosed later than men, delaying the initiation of lipid-lowering therapies [8]. In primary care settings, women are 22% less likely than men to achieve LDL-C targets within 180 days of starting or adjusting LLT, regardless of therapy intensity, cardiovascular risk category, age, mental health status, or socioeconomic factors [9]. This disparity may contribute to poorer cardiovascular outcomes in women.

Postmenopausal women experience a significant increase in LDL-C levels, attributed to decreased estrogen, which adversely affects lipid profiles [10]. This elevation in LDL-C heightens the risk of developing comorbid conditions such as hypertension, type 2 diabetes, and metabolic syndrome [11]. Additionally, women with hypercholesterolemia are at increased risk for osteoporosis and certain cancers, including breast cancer, due to complex interactions between lipid metabolism and hormonal changes [11].

Aging presents unique health challenges for both men and women, but their disease trajectories, the onset of comorbidities, and their progression may differ between genders. Generally, women tend to live longer than men but also face more comorbidities and a higher incidence of chronic diseases in old age. However, there are knowledge gaps regarding how biological, hormonal, and social factors influence these health differences between men and women as they age [3].

Real-world data (RWD) can facilitate the investigation of these differences, as traditional clinical trials often fail to capture the diversity of the population or reflect the daily experiences of men and women [12, 13]. Andalusia is the largest region in Spain and the third largest in Europe, with a population of 8.5 million, comparable to countries like Switzerland and Austria. The Andalusian Health System has been collecting medical data from their used since 2001 in the Population Health Database (BPS, Spanish acronym for *Base Poblacional de Salud*) [14].

This study leverages the vast amount of RWD available within the BPS, where 557,034 patients diagnosed with hypercholesterolemia, between 1^st^ January 2016 and 31^st^ December 2022, were found. By utilizing this extensive dataset, the study aims to describe and quantify the impact of hormonal transitions, particularly menopause, on women’s health by comparing aging and comorbidities in men and women before and after the menopause age, aiming to provide a more accurate understanding of gender differences in the progression of chronic diseases. The findings reveal a remarkable difference between men and woman both, in the evolution of the disease and the associated comorbidities. These results will be useful for future clinical recommendations and will contribute to improve the management of the patients, ultimately enhancing their quality of life. This research also underscores the transformative potential of RWD [15] in refining healthcare practices and guiding decision-making in patient care.

## Material and Methods

### Data

The Andalusian Biomedical Research Ethics Coordinating Committee approved the study entitled: “Retrospective observational study to determine the characteristics and treatment patterns in patients with atherosclerotic cardiovascular disease and familial hypercholesterolemia in Andalusia. “ (March 28, 2023, Minutes 03/23) and waived informed consent for the secondary use of clinical data for research purposes.

The target population of the study were adult Andalusian patients with a diagnosis of hypercholesterolemia (ICD-10: E78.0, E78.00, E78.01, E78.2, E78.49) for whom data are available in BPS during the period between 1st January 2016 and 31st December 2022. Clinical data, along with comorbidities of different categories (Cardiovascular, Dermatological, Bone and Joint, Neurological, Renal and Urinary, Mental Health and Substance Use, Liver, Ocular, Cancers, Endocrine and Metabolic, Gastrointestinal, Collagen disease and vasculitis, Cerebrovascular, Respiratory, Congenital and Developmental Disorders, Infectious), obesity, current smoker status and alcohol consumption, were used (See Supplementary Table 1 for a detailed list of the comorbidities used).

### Design of the study

This is a retrospective, multicenter, non-interventional, retrospective observational cohort study of patients diagnosed with hypercholesterolemia using data included in the BPS.

The first diagnosis of hypercholesterolemia (index episode) in the retrospective cohort will mark the starting point of follow-up of the individual within the cohort. The follow-up period will be considered completed in the event of death, loss to follow-up, or termination of the study.

The study period covers from January 1, 2016 to December 31, 2022. Inclusion and exclusion criteria will be applied at the patient’s index date.

Inclusion criteria are: patients ≥ 18 years, with a hypercholesterolemia diagnosis (ICD-10 codes: E78.0, E78.00, E78.01), and regular follow-up guaranteed for, at least, 1 annual health record during the study period. Patients who at the index were in the palliative service, or with code Z51.5 Contact for palliative care.

### Statistical methods

To identify differences between women and men, categorical variables were presented as frequencies and percentages. Continuous variables were summarized using means and standard deviations (SD) or medians and interquartile ranges (IQR), depending on the data distribution. The X^2^ or Fisher’s exact test was used to compare categorical variables, while the t-test or Wilcoxon rank-sum test was used to compare continuous variables, depending on their distribution. The Kruskal–Wallis test was used to compare cost differences between more than two groups. All analyses were two-sided, and statistical significance was set at p < 0.05.

To further investigate the relationship between the number of diagnoses, age, and sex, an analysis of the odds ratio trajectories was conducted. The data were stratified by age and sex, and the number of diagnoses within each stratum was determined. For each age, the odds of diagnosis in women relative to men was calculated. Confidence intervals (95%) for each odds ratio were constructed using the normal approximation.

The analysis utilized statistical methods to investigate trends in disease prevalence across sex and age. Each disease trajectory was modeled after the logarithmic odds ratios, reflecting the relative prevalence of diseases between women and men across the 35 - 65 age range. K-means clustering was employed to group diseases with similar log odds ratio trajectories over age, with the optimal number of clusters determined using the elbow method [16].

The statistical analysis utilized Python (version 3.10.12) and open-source libraries, including Numpy (version 2.1.1) [17], SciPy (version 1.14.1) [18], statsmodels (version 0.14.3) [19] and TableOne package (version 0.9.1) [20]. Trajectory clustering was done using scikit-learn (version 1.5) [21], while kneed (version 0.8.5) [22] was used to compute the optimal number of clustering trajectories. Data visualizations, such as boxplots and bar charts, were generated using Seaborn (version 0.13.2) [23].

### Sample size estimation

In conventional studies using hospital databases, the size calculation underlies the need to obtain a portion or subset of individuals with specific characteristics (i.e. sample) that is representative of the whole, and to be able to respond to the study objectives with adequate power and precision so that the results can be generalized to the whole population. This study uses Real World Data (RWD) and the major difference in this approach is precisely that this study allows the inclusion of all patients who meet the selection criteria, i.e. the sample is directly the entire target population of hypercholesterolemia patients in Andalusia. However, an assessment is made of the sample size required for the study.

The prevalence of hypercholesterolemia in the Spanish adult population has been estimated at 50% [24]. With this prevalence, the minimum required sample size would be 1187 for the margin of error or absolute precision of 3% in estimating the prevalence with 95% confidence and considering the potential loss/attrition of 10%. The anticipated 95% CI is (47%, 53%). This minimum sample size is calculated using the ScalaR SP [25]. The sample used in this study far exceeds the minimum required sample size.

### Secure data management

The data management circuit was designed to minimize the relative risk as described in the Impact Assessment on Data Protection analysis [26] and following the regulation for the use of medical data for research purposes in Andalusia (Joint Resolution 1/2021 of the General Secretariat for Research, Development and Innovation in Health of the Regional Ministry of Health and Families and the Management Directorate of the Andalusian Health Service [27]). Briefly, the data corresponding to the study approved by the ethics committee is requested to BPS. There, the data is extracted and pseudonymized by BPS personnel and then transferred to the iRWD [28, 29], a Secure Processing Environment (SPE) specifically designed for the secure analysis of data protected by the GDPR, being compliant with the definition of SPE as described in Article 50 of the Resolution of the European Parliament of 24 April 2024 on the proposal for a Regulation of the European Parliament and of the Council on the EHDS [30].

## Results

### Cohort of study

A total of 557,034 patients diagnosed with hypercholesterolemia meeting the inclusion criteria between 1st January 2016 and 31st December 2022 were found. This cohort is composed of 227,834 men and 329,200 women, and the average age (and standard deviation) of hypercholesterolemia diagnosis was 56.0 (13.2) and 59.1 (13.2) years old, respectively. Table 1 Summarizes the basic data of the cohort studied, including the different comorbidity categories (see Supplementary Table 1 for the whole list of comorbidities).

**Table 1.** Basic data of the cohort studied, along with the comorbidities by categories, with the mean age at diagnosis with the [Q1,Q3] interquartile range, and the number of individuals diagnosed with this comorbidity. Differences between men and woman are tested for ages with a Kruskal-Wallis test and for number of affected by a X2 test.

| Comorbidity category | Diagnosis | Men | Women | P-Value |
| --- | --- | --- | --- | --- |
| **Cardiovascular Diseases** | Age [Q1,Q3] | 57.0 [49.0,65.0] | 59.0 [51.3,68.0] | <0.001 |
|  | N(%) | 141374 (62.3) | 196303 (59.9) | <0.001 |
| **Dermatological Disorders** | Age [Q1,Q3] | 52.0 [42.0,61.0] | 54.0 [45.0,62.0] | <0.001 |
|  | N(%) | 12445 (5.5) | 16432 (5.0) | <0.001 |
| **Bone and Joint Disorders** | Age [Q1,Q3] | 59.0 [51.0,66.0] | 59.0 [52.0,66.3] | <0.001 |
|  | N(%) | 89328 (39.4) | 205724 (62.8) | <0.001 |
| **Neurological Disorders** | Age [Q1,Q3] | 65.0 [54.0,74.9] | 70.0 [58.0,78.0] | <0.001 |
|  | N(%) | 26159 (11.5) | 40214 (12.3) | <0.001 |
| **Renal and Urinary Disorders** | Age [Q1,Q3] | 57.0 [47.0,68.0] | 58.0 [48.0,70.0] | <0.001 |
|  | N(%) | 47767 (21.0) | 67637 (20.6) | <0.001 |
| **Mental Health and Substance Use** | Age [Q1,Q3] | 52.0 [43.0,62.0] | 54.0 [45.5,64.0] | <0.001 |
|  | N(%) | 112541 (49.6) | 194227 (59.3) | <0.001 |
| **Liver Diseases** | Age [Q1,Q3] | 55.0 [47.0,64.0] | 60.0 [53.0,67.5] | <0.001 |
|  | N(%) | 31180 (13.7) | 35140 (10.7) | <0.001 |
| **Ocular Diseases** | Age [Q1,Q3] | 63.0 [54.0,72.0] | 65.2 [57.0,74.0] | <0.001 |
|  | N(%) | 24819 (10.9) | 40048 (12.2) | <0.001 |
| **Cancers** | Age [Q1,Q3] | 65.0 [57.0,73.0] | 64.0 [55.0,73.0] | <0.001 |
|  | N(%) | 15374 (6.8) | 15345 (4.7) | <0.001 |
| **Endocrine and Metabolic Disorders** | Age [Q1,Q3] | 52.0 [44.0,61.0] | 56.0 [48.0,65.0] | <0.001 |
|  | N(%) | 145737 (64.2) | 191320 (58.4) | <0.001 |
| **Gastrointestinal Disorders** | Age [Q1,Q3] | 54.0 [45.0,64.0] | 58.0 [50.0,67.0] | <0.001 |
|  | N(%) | 25635 (11.3) | 50344 (15.4) | <0.001 |
| **Collagen disease and vasculitis** | Age [Q1,Q3] | 64.0 [53.0,73.0] | 61.0 [53.0,71.0] | <0.001 |
|  | N(%) | 3107 (1.4) | 11820 (3.6) | <0.001 |
| **Cerebrovascular Disorders** | Age [Q1,Q3] | 68.0 [60.5,76.0] | 71.0 [62.0,79.0] | <0.001 |
|  | N(%) | 7022 (3.1) | 6321 (1.9) | <0.001 |
| **Respiratory Diseases** | Age [Q1,Q3] | 58.0 [47.0,67.0] | 56.0 [46.0,66.0] | <0.001 |
|  | N(%) | 42893 (18.9) | 56651 (17.3) | <0.001 |
| **Congenital and Developmental Disorders** | Age [Q1,Q3] | 57.0 [48.0,66.0] | 59.0 [47.0,70.0] | <0.001 |
|  | N(%) | 1484 (0.7) | 1552 (0.5) | <0.001 |
| **Infectious Diseases** | Age [Q1,Q3] | 42.0 [36.0,49.0] | 41.0 [35.0,49.0] | 0.055 |
|  | N(%) | 1399 (0.6) | 590 (0.2) | <0.001 |
| **Total patients** |  | 226,962 | 327,710 |  |

As expected, differences between sex and age onset are significant for all the comorbidity types, except for the case of age onset in infectious diseases. Supplementary Table 2 contains the detailed list of comorbidities for each disease category.

### Influence of sex in the hypercholesterolemia diagnosis

Figure 1 illustrates the distribution of hypercholesterolemia diagnoses across age groups, separated by sex. Several key patterns emerge from this data that provide insights into the prevalence and progression of this condition. Firstly, an age-related pattern steady increase of hypercholesterolemia diagnoses with age in both sexes is observed, peaking between 55 and 60 years, followed by a gradual decline in older populations. The bell-shaped curve is consistent with age-related metabolic changes, dietary patterns, and increased screening in middle-aged individuals.

**Figure 1.**
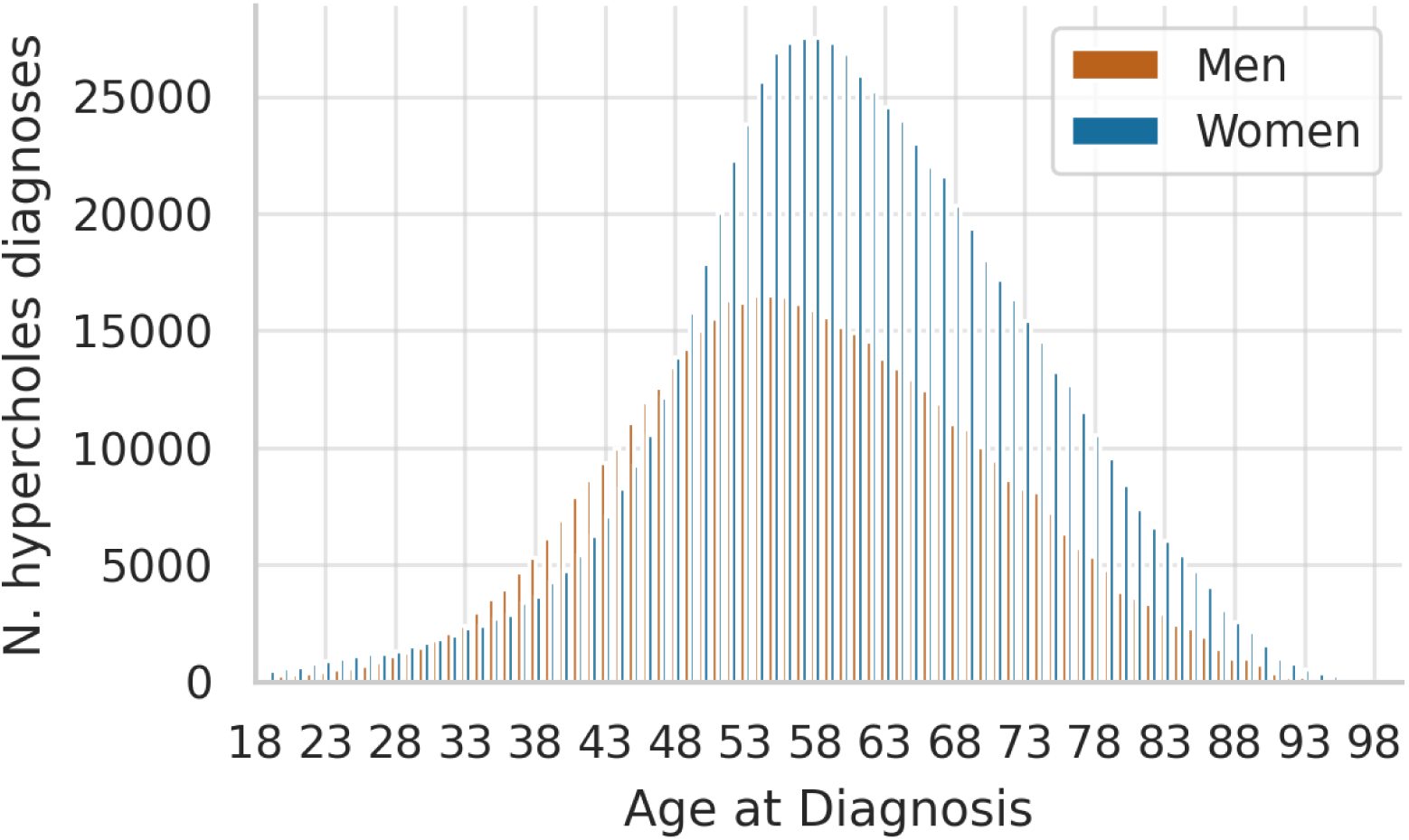
Number of hypercholesterolemia diagnoses in the cohort under study across ages, stratified by sex (men and women).

However, sex-specific patterns are dominant, with diagnoses in men increasing more rapidly at younger ages (30–50 years), surpassing those of women in early adulthood. The incidence in men peaks slightly earlier (around 50–55 years), suggesting that hypercholesterolemia may manifest earlier in males due to factors such as lifestyle, diet, or inherent biological differences (e.g., higher baseline LDL levels). Diagnoses in women increase more gradually but surpass those of men around 50–55 years and remain higher in older age groups. This trend aligns with the postmenopausal phase, during which estrogen levels decline, leading to adverse lipid profile changes, including higher LDL and total cholesterol. The sharp rise in women’s hypercholesterolemia diagnoses around the age of 50 coincides with the typical onset of menopause. This strongly supports the well-documented relationship between reduced estrogen levels and worsening cholesterol profiles. After menopause, women show consistently higher rates of hypercholesterolemia than men, reflecting long-term cardiovascular risk. Beyond the age of 70, the incidence of diagnoses declines for both sexes, likely due to survivorship bias (healthier individuals are overrepresented in older cohorts) or reduced screening frequency.

### Influence of sex in the comorbidity frequency

Figure 2 illustrates the distribution of diagnosed comorbidities across age ranges, stratified by sex (men and women). The general trend indicates a clear age-related increase in the number of comorbidities, peaking in middle age and gradually declining in older age groups. There are some notable differences between men and women on menopause and sex-based health disparities.

**Figure 2.**
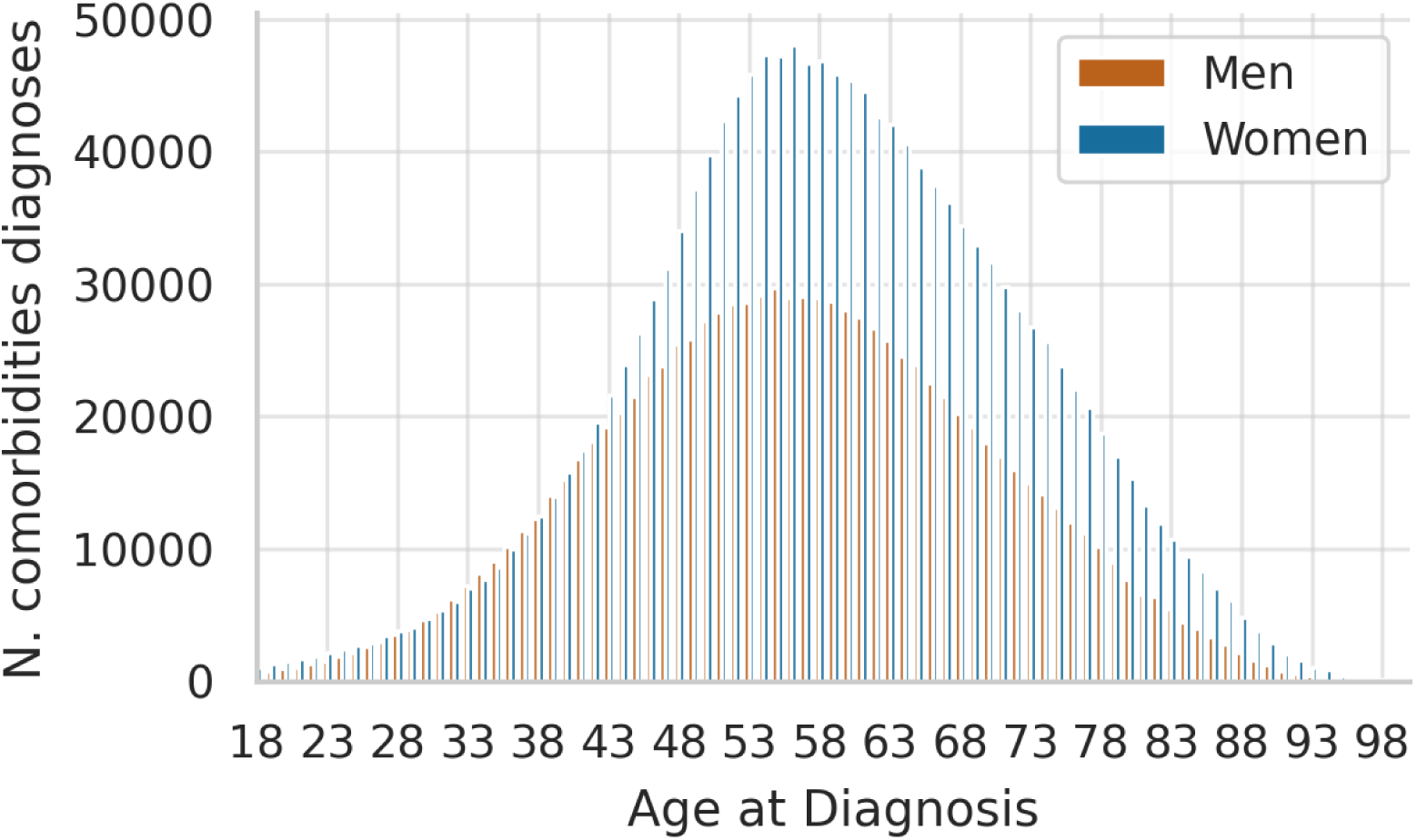
Number of comorbidities diagnosed in the cohort under study across ages, stratified by sex (men and women).

As an overall trend both sexes exhibit an increase in the number of comorbidities with age. The peak incidence of comorbidities occurs around the age of 55–60 for both men and women. However, there are remarkable sex-specific differences: women consistently display a higher incidence of comorbidities across nearly all age ranges, particularly between the ages of 45 and 70. The difference becomes particularly pronounced around the menopausal transition period (ages 45–55), suggesting that hormonal changes associated with menopause might influence the onset or progression of comorbid conditions. After menopause (typically 50+ years), women continue to exhibit higher rates of comorbidities than men. This might be linked to physiological and metabolic changes post-menopause, such as increased cardiovascular risk, changes in bone density, or alterations in immune function. In older age groups (75+ years), the gap between men and women narrows, with a slight decline in comorbidity incidence in both sexes. This might be influenced by survivor effects or a plateau in new diagnoses.

### Sex-related comorbidity pattern in the post-menopause age

In order to understand the effect of menopause in the occurrence of different comorbidities the proportion of diagnoses in women versus men is assessed. Figure 3 portraits the odd ratios of the total occurrence of comorbidities in woman versus man, in which a significant trend of increase in woman morbidities with respect to man is observed immediately after the 50 years. This global trend is followed to some extent by many of the comorbidities studied.

**Figure 3.**
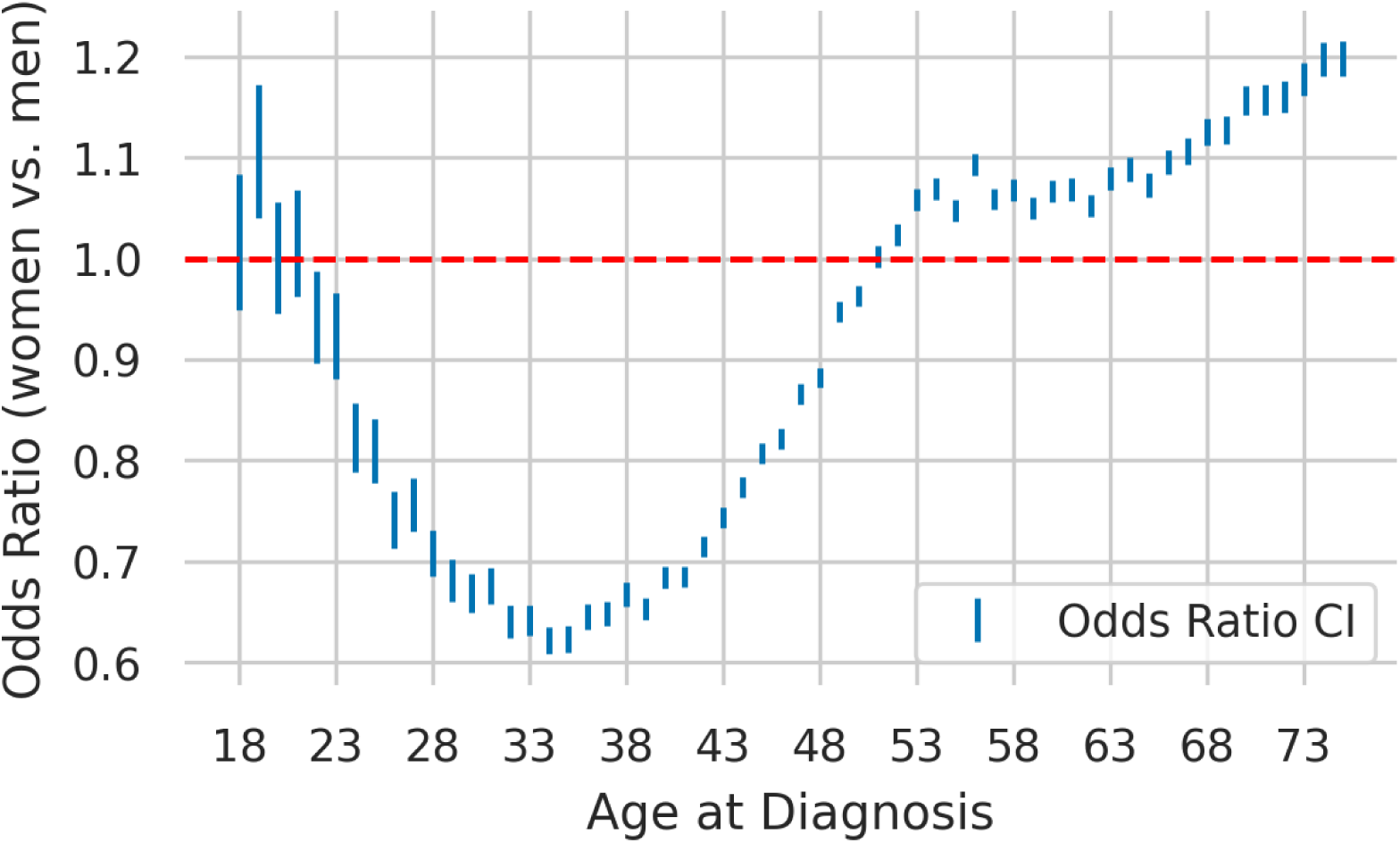
Odd ratios and confidence intervals (CI) of the total occurrence of comorbidities in woman versus man.

When individual odds ratio trajectories for various morbidities are analyzed using K-means clustering (see Methods), two primary trends emerge, as depicted in Figure 4.

**Figure 4.**
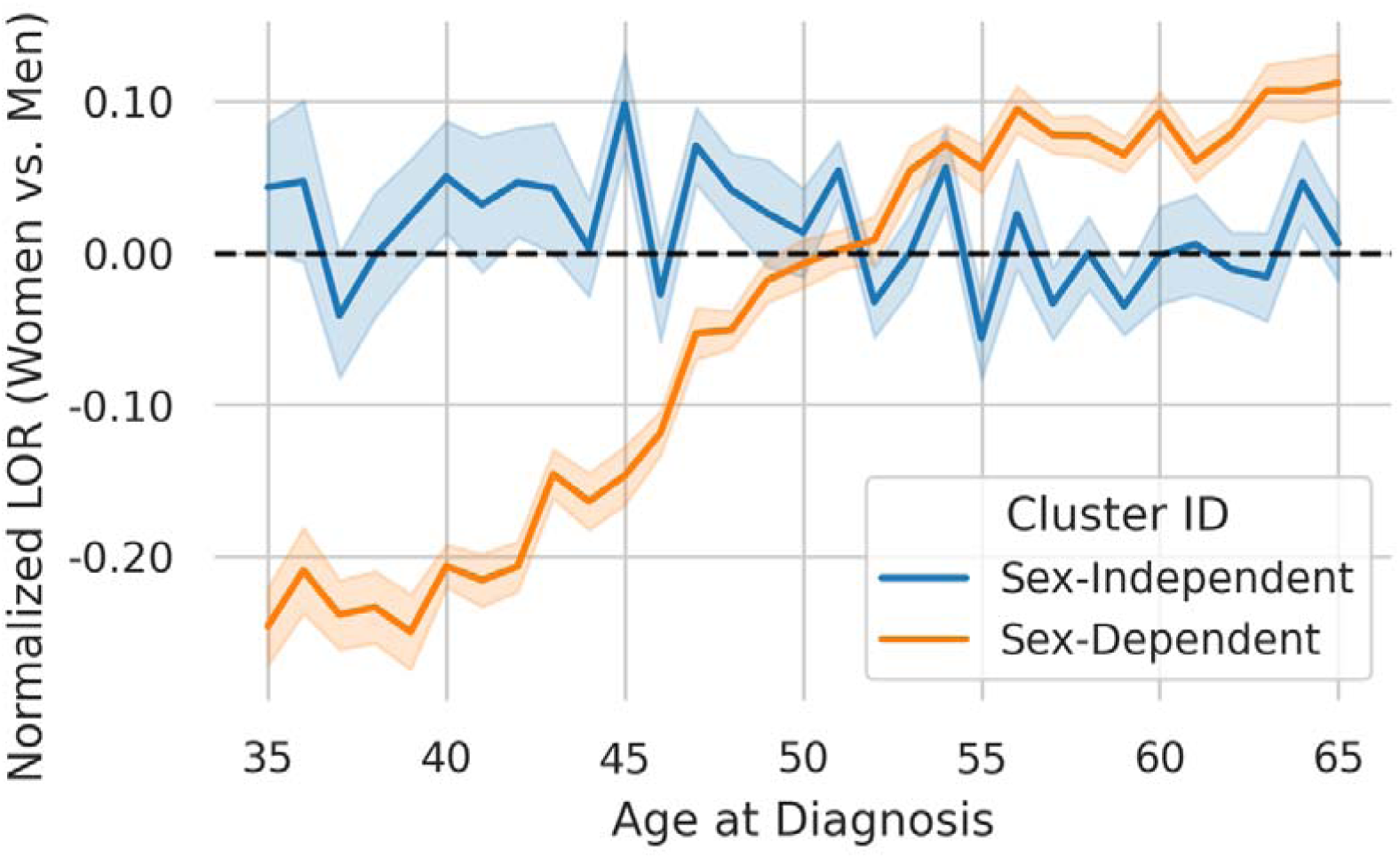
Adjusted log-OR of the occurrences of comorbidities in woman versus man. Two clusters of diseases are represented.

**Figure 5.**
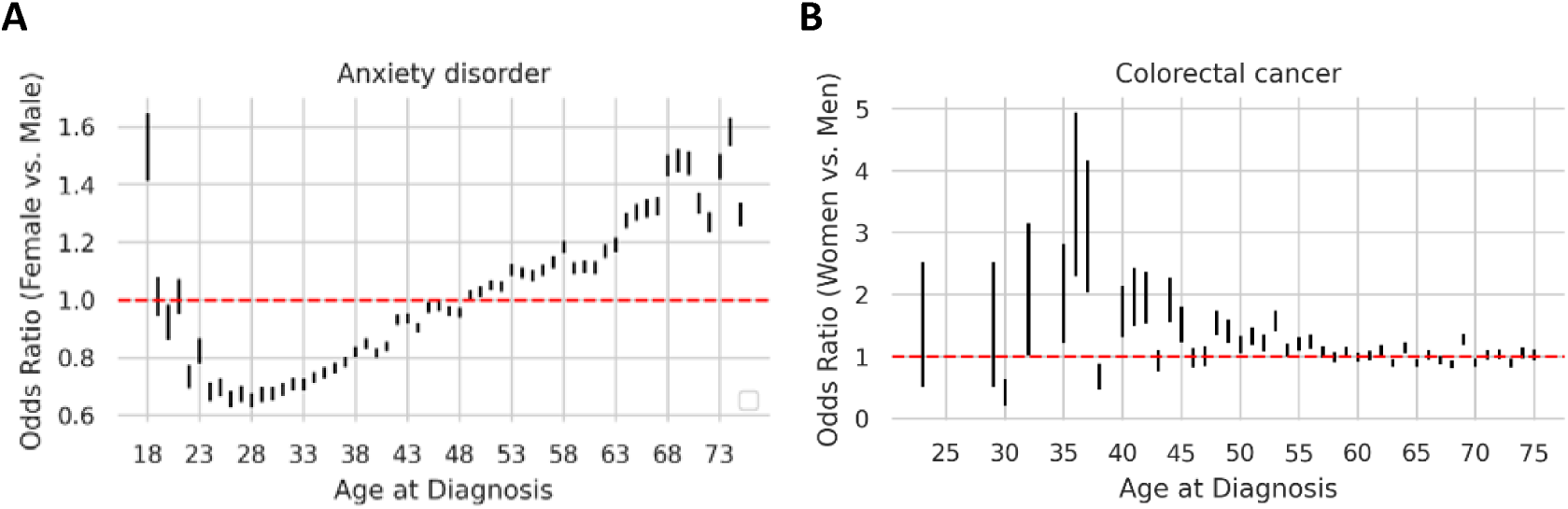
OR trajectories for Anxiety disorder, a sex-dependent mental disorder (left) and colorectal cancer, a sex-independent disorder (right).

One cluster predominantly includes sex-dependent conditions, encompassing mental, cardiovascular, endocrine, metabolic, bone and joint, neurological, and liver disorders. The other group, includes sex-independent diseases, some of them age-related, such as cancers (see Figure 4B) or intellectual disability, and other related to genetic factors, such as congenital cardiac and circulatory anomaly or infectious, such as HIV.

Supplementary Table 3 contains a detailed description of the diseases following both trends depicted in Figure 4.

Table 1, lists the most frequent comorbidities (those accounting for more than 1.5% of the patients with one or more diagnoses) observed in the cohort under study (a complete list is available in Supplementary Table 4).

**Table 1.** The most frequent comorbidities in woman. Number of patients with at least one comorbidity (second and third columns represent observations in patients younger than 50 years, and fourth and fifth columns in patients 50 years old and older). A X^2^ test has been conducted to detect significant increases of comorbidity diagnoses in women with respect to men when the older (post-menopausal age) are compared to the younger, and the sixth column reports the p-value. The seventh column represent the total incidence ratio woman/man between the respective increase in comorbidities in older with respect to younger. The last column is the percentage of woman with the comorbidity with respect to all women with comorbidities.

| Comorbidities | <50 |  | ≥50 |  | P-Value | Ratio | % |
| --- | --- | --- | --- | --- | --- | --- | --- |
|  | Men | Women | Men | Women |  |  |  |
| Osteoarthritis, spondylosis | 14,247 | 28,405 | 66,032 | 156,067 | <0.001 | 1.19 | 12.90 |
| Hypertension | 39,138 | 38,776 | 91,532 | 147,538 | <0.001 | 1.63 | 12.20 |
| Anxiety disorder | 29,695 | 64,875 | 31,374 | 94,916 | <0.001 | 1.38 | 7.85 |
| Hypothyroidism | 5,577 | 28,855 | 10,161 | 57,628 | <0.001 | 1.10 | 4.76 |
| Diabetes | 14,358 | 10,845 | 44,874 | 53,588 | <0.001 | 1.58 | 4.43 |
| Mixed hyperlipidemia | 41,025 | 22,744 | 34,207 | 45,362 | <0.001 | 2.39 | 3.75 |
| Osteoporosis | 322 | 2,081 | 2,623 | 42,255 | <0.001 | 2.49 | 3.49 |
| Mood disorder | 6,295 | 14,456 | 10,798 | 41,216 | <0.001 | 1.66 | 3.41 |
| Obesity | 13,278 | 15,701 | 22,772 | 36,240 | <0.001 | 1.35 | 3.00 |
| Urinary stones | 14,128 | 18,296 | 21,030 | 34,946 | <0.001 | 1.28 | 2.89 |
| Gastroesophageal reflux disease (GERD) | 7,384 | 7,999 | 12,483 | 30,642 | <0.001 | 2.27 | 2.53 |
| Tobacco dependence | 22,162 | 18,437 | 28,919 | 23,832 | 0.48 | 0.99 | 1.97 |
| Asthma | 9,922 | 16,714 | 7,934 | 23,426 | <0.001 | 1.75 | 1.94 |
| Other arthropathy | 5,452 | 5,943 | 13,133 | 23,126 | <0.001 | 1.62 | 1.91 |
| Heart failure | 1,138 | 711 | 16,412 | 22,915 | <0.001 | 2.23 | 1.89 |
| Chronic Obstructive Pulmonary Disease | 4,426 | 3,929 | 25,492 | 22,087 | 0.313 | 0.98 | 1.83 |
| Hepatic steatosis (Fatty liver) | 7,472 | 4,014 | 15,307 | 21,695 | <0.001 | 2.64 | 1.79 |
| Pure hyperglyceridemia | 24,515 | 11,339 | 18,128 | 20,303 | <0.001 | 2.42 | 1.68 |
| Fibromyalgia | 558 | 10,749 | 823 | 20,035 | <0.001 | 1.26 | 1.66 |
| Glaucoma | 1,932 | 2,861 | 9,424 | 19,129 | <0.001 | 1.37 | 1.58 |
| Atrial fibrillation | 1,268 | 514 | 15,073 | 18,787 | <0.001 | 3.07 | 1.55 |

The data highlights that the prevalence of comorbidities increases with age in both sexes, but this rise is disproportionately higher in women post-menopause and in most of the cases statistically significant. Mental and behavioral disorders, such as anxiety, other organic mental disorders and mood disorders, consistently exhibit higher prevalence in women, with notable post-menopausal increases in conditions like dementia. Cardiovascular diseases also show marked post-menopausal increases, with heart failure, Atrial fibrillation, peripheral arterial disease and ischemic heart disease highlighting the loss of estrogen’s protective effects. Bone-related conditions, such as osteoarthritis and spondylosis, osteoporosis, along with metabolic disorders like pure hyperglyceridemia, obesity or metabolic syndrome further underscore the significant impact of menopause on women’s health.

Among cancers, most conditions exhibit a significant age-related increase in both sexes, as anticipated, with few exceptions showing sex-specific differences. Supplementary Table 4 highlights that Non-Hodgkin lymphoma (p = 0.01) and leukemia (p = 0.003) have statistically significant higher prevalence in women compared to men post-menopause. Additionally, marginally significant differences are noted in bladder cancer (p = 0.031) and bronchus and lung cancer (p = 0.044).

Table 2 and the OR plots in the Supplementary Figure 1 illustrate the pronounced disparities in comorbidities between men and women in the post-menopausal period. Conditions like gout and other crystal-induced arthropathies (woman to man incidence ratio, WMIR= 4.65), with a significant Odd-ratio (OR) over 60 years in woman (see Supplementary Figure AD) as well as other organic mental disorders (WMIR=4.02), with a significant OR in women over 70 years (Supplementary Figure BH), followed by another mental condition, autism spectrum disorder (WMIR=3.44) exhibit the largest disparities, revealing that post-menopausal women face greater relative increases in these conditions compared to men. In general, for mental disorders notable ratios are observed, like schizophrenic disorder (WMIR=2.73), which presents a significant OR in women over 50 years (Supplementary Figure BQ) and dependence on other substances (WMIR=2.24), with also a significant OR in women over 50 years (Supplementary Figure V) indicating that hormonal or neurochemical changes post-menopause may significantly impact mental health in women and underscoring a heightened vulnerability among women in older age. Among cardiovascular diseases, atrial fibrillation (WMIR=3.07) and ischemic heart disease (WMIR=2.57) show substantial increases in women, likely due to the loss of estrogen’s protective effects on cardiac and vascular systems Both diseases present a significant OR but at older ages, over the 65 years (Supplementary Figures I and AP, respectively). For metabolic and lipid-related conditions, hepatic steatosis (WMIR=2.74), pure hyperglyceridemia (WMIR=2.42), and mixed hyperlipidemia (WMIR=2.39), with a significant OR soon after the menopause period (Supplementary Figures AH, BM and AW, respectively), highlight the adverse effects of hormonal changes on metabolic health in post-menopausal women. Notably, osteoporosis (WMIR=2.49) also with a significant OR soon after the menopause period (Supplementary Figure BD), reflects the critical impact of estrogen loss on bone health, with post-menopausal women disproportionately affected. Overall, these findings emphasize the need for sex-specific prevention and management strategies for women’s health post-menopause.

**Table 2.** Diagnoses with most pronounced frequency increase on post-menopausal women with respect to men.

| Comorbidities | <50 |  | ≥50 |  | Ratio | p-value |
| --- | --- | --- | --- | --- | --- | --- |
|  | Men | Women | Men | Women |  |  |
| Gout and other crystal-induced arthropathies | 6,607 | 515 | 17,065 | 6,180 | 4.65 | <0.001 |
| Other organic mental disorder | 1,435 | 670 | 8,531 | 16,016 | 4.02 | <0.001 |
| Autism spectrum disorder | 71 | 25 | 33 | 40 | 3.44 | <0.001 |
| Dependence on other substances | 2,502 | 758 | 1,295 | 1,343 | 3.42 | <0.001 |
| Atrial fibrillation | 1,268 | 514 | 15,073 | 18,787 | 3.07 | <0.001 |
| Schizophrenic disorder | 2,294 | 1,510 | 1,235 | 2,217 | 2.73 | <0.001 |
| Hepatic steatosis (Fatty liver) | 7,472 | 4,014 | 15,307 | 21,695 | 2.64 | <0.001 |
| Ischemic heart disease | 3,242 | 789 | 20,783 | 13,022 | 2.57 | <0.001 |
| Osteoporosis | 322 | 2,081 | 2,623 | 42,255 | 2.49 | <0.001 |
| Pure hyperglyceridemia | 24,515 | 11,339 | 18,128 | 20,303 | 2.42 | <0.001 |
| **Mixed hyperlipidemia** | 41,025 | 22,744 | 34,207 | 45,362 | 2.39 | <0.001 |
| **Dementia** | 107 | 118 | 5,058 | 13,168 | 2.36 | <0.001 |

## Discussion

For clinicians, acknowledging sex-specific and gender-specific factors can improve diagnosis, treatment, and disease management. Research has consistently shown that sex and gender play significant roles in the onset, progression, and outcomes of diseases. However, these insights have yet to be systematically integrated into everyday clinical practice. To achieve so, this study presents a detailed characterization large cohort of 557,034 patients, a dataset far larger than most clinical trials, that describes in detail the impact of sex and age in hypercholesterolemia and its associated comorbidities. The region-wide scale of the study ensures the inclusion of diverse patient profiles, reflecting real-world variability in disease presentation and management, emphasizing the transformative potential of RWD in understanding complex biomedical problems [13, 31]. Compared to controlled clinical trials, RWD offers a more nuanced understanding of health conditions as they occur in everyday clinical settings, a key advantage that has been increasingly recognized in epidemiological research [12].

The findings align with existing knowledge regarding hypercholesterolemia’s sex-specific nature. Women were older at diagnosis than men, consistent with studies highlighting delayed diagnosis in women due to less aggressive LDL-C screening and treatment initiation [8, 10]. The longer life expectancy of women likely contributes to the observed disparities, as older age is associated with greater cardiovascular risk and multimorbidity [3]. The higher mortality age in women further underscores the significance of understanding sex-specific disease progression.

Moreover, this dataset confirms previously established patterns, such as the sharper rise in hypercholesterolemia diagnoses among women post-menopause. This finding correlates with reduced estrogen levels, which adversely affect lipid profiles and cardiovascular health [10]. By incorporating real-world variability, these results extend existing knowledge, reinforcing the importance of menopause in disease progression.

The analysis of comorbidity frequency highlights significant sex-based differences, particularly during and after the menopausal transition. Women consistently displayed higher rates of comorbidities than men, especially in the 45–70 age range, which coincides with menopause. This period is marked by hormonal changes, including a dramatic reduction in estrogen, which has protective effects on cardiovascular health and metabolic regulation [10]. The study findings align with prior research showing increased risks of hypertension, osteoporosis, and type 2 diabetes in postmenopausal women compared to men [32]. Also, menopause significantly influences the development of metabolic syndrome in women. The transition to menopause is associated with increased central adiposity, adverse lipid profiles, and insulin resistance, collectively elevating the risk of metabolic syndrome. Postmenopausal women exhibit higher rates of metabolic syndrome compared to their premenopausal counterparts, independent of aging effects [33]. This heightened prevalence contributes to an increased risk of cardiovascular diseases and type 2 diabetes in postmenopausal women [34].

Notably, the gap in comorbidity prevalence narrows after the age of 75, likely due to survivorship bias or reduced diagnostic intensity in older populations. These trends underscore the importance of considering sex and age in disease prevention and management strategies. For instance, targeted interventions, such as early screening and tailored therapeutic strategies, could mitigate the elevated comorbidity burden in postmenopausal women [35]. Moreover, the higher prevalence of mental health disorders, such as anxiety and mood disorders, among women across all age groups, and the sharp increase after menopause, suggests the need for integrated care approaches that address both physical and mental health conditions [36].

Women generally live longer than men, but this increased lifespan is accompanied by a higher incidence of chronic diseases and comorbidities in later years. While men may experience earlier mortality, women are more likely to suffer from conditions such as osteoporosis, arthritis, and cardiovascular diseases. Understanding why these differences occur is crucial, yet traditional clinical research has largely ignored the interplay of biological, hormonal, and social factors influencing aging [3, 37].

Hormones play a pivotal role in women’s health. Estrogen, for example, not only regulates reproduction but also protects against several chronic conditions, such as heart disease and osteoporosis. As women age, the transition to menopause—a period characterized by a significant drop in estrogen levels—triggers physiological changes that can affect their overall health and quality of life. These changes extend beyond typical menopausal symptoms like hot flashes and vaginal dryness, potentially leading to increased cardiovascular risk, cognitive decline, and bone density loss [38–41]. Clinicians should consider incorporating sex and gender in their decision-making to practice precision medicine that integrates fundamental components of patient individuality. Recognizing the biological and environmental factors that affect the disease course is imperative to optimizing care for each patient. Research highlights the myriad ways sex and gender play a role in health and disease. However, these clinically relevant insights have yet to be systematically incorporated into care [42].

The results presented here underscore the critical role of lack of estrogens in menopause in the pathophysiology of hypercholesterolemia in women. The earlier and steeper rise in men suggests sex-specific risk factors that should be investigated further, such as behavioral patterns or genetic predispositions. Targeted interventions, such as lifestyle modifications or earlier lipid screening in men and focused management of lipid profiles in postmenopausal women, may be warranted. Additionally, the pronounced sex differences in older adults highlight the need for age- and sex-specific approaches to hypercholesterolemia and other comorbidities treatment and prevention. It has been described previously, the null effect of statin therapy on primary CVD prevention in women contrasts with that of men. With careful separation of women from men and primary from secondary prevention trials, the first sex-specific meta-analysis to analyze primary CVD prevention trials showed that statin therapy reduced CHD in men but not in women [42]. In this regard, Hormone Replacement Therapy has been proposed a sex-specific and time-dependent primary CVD prevention therapy that concomitantly reduces all-cause mortality, as well as other aging-related diseases with an excellent risk profile [42]. Keeping in mind that prevention strategies must be personalized, health care providers and patients can use cumulated HRT data in making clinical decisions concerning chronic disease prevention including CVD and mortality reduction.

Currently, many women, suffering from conditions ranging from cancer to anxiety, see their health issues mistakenly addressed from a male perspective. The utilization of a large RWD cohort in our study allows for a more accurate and detailed understanding of these sex-specific differences, providing a robust foundation for developing tailored interventions. By capturing a wide array of patient experiences and outcomes, RWD facilitates the identification of patterns that inform clinical decision-making and health policy, ultimately contributing to improved patient care [12, 13, 31].

## Data Availability

The datasets analyzed during the current study are available in the Population Health Database repository (https://www.sspa.juntadeandalucia.es/servicioandaluzdesalud/profesionales/sistemas-de-informacion/base-poblacional-de-salud), subject to controlled access according to the regulation for the use of medical data for research in the Andalusian Health System (https://www.sspa.juntadeandalucia.es/servicioandaluzdesalud/sites/default/files/sincfiles/wsas-media-sas_normativa_mediafile/2021/resolucion_conjunta_acceso_a_datos_investigacion_def_20211201%28F%29.pdf).

## Acknowledgements

The infrastructure where this work has been carried out has been funded by the Consejeria de Salud y Consumo, Junta de Andalucía (IE19_259 FPS). We acknowledge the help of Dolores Muñoyerro-Muñiz, Román Villegas, and other BPS curators on issues related with the complexity of clinical data management and interpretation.

