## Supplementary Table 1 for "The Impact of Menopause on Hypercholesterolemia and Comorbidities: A Population-Based Study"

**Supplementary Table 1**. Basic data of the cohort studied, along with the comorbidities, with the mean age at diagnosis with the [Q1,Q3] interquartile range, and the number of individuals diagnosed with this comorbidity. Differences between men and woman are tested for ages with a Kruskal-Wallis test and for number of affected by a X2 test.

|  |  | **Men** | **Women** | **P-Value** |
| --- | --- | --- | --- | --- |
| **Disease** | **Total n** | 226962 | 327710 |  |
| **Hypertension** | **Age diagnosis, median [Q1,Q3]** | 56.0 [48.0,64.0] | 58.0 [51.0,66.0] | <0.001 |
|  | **n(%)** | 130670 (57.6) | 186314 (56.9) | <0.001 |
| **Psoriasis** | **Age diagnosis, median [Q1,Q3]** | 52.0 [42.0,61.0] | 54.0 [45.0,62.0] | <0.001 |
|  | **n(%)** | 10651 (4.7) | 12445 (3.8) | <0.001 |
| **Osteoporosis** | **Age diagnosis, median [Q1,Q3]** | 65.0 [57.0,74.0] | 64.0 [57.0,71.0] | <0.001 |
|  | **n(%)** | 2945 (1.3) | 44336 (13.5) | <0.001 |
| **Osteoarthritis, spondylosis** | **Age diagnosis, median [Q1,Q3]** | 59.0 [52.0,67.0] | 59.0 [53.0,67.0] | 0.004 |
|  | **n(%)** | 80279 (35.4) | 184472 (56.3) | <0.001 |
| **Extrapyramidal syndrome** | **Age diagnosis, median [Q1,Q3]** | 66.0 [57.0,74.0] | 69.0 [61.0,76.0] | <0.001 |
|  | **n(%)** | 4316 (1.9) | 8088 (2.5) | <0.001 |
| **Chronic renal insufficiency** | **Age diagnosis, median [Q1,Q3]** | 70.0 [61.0,78.0] | 76.0 [67.0,82.0] | <0.001 |
|  | **n(%)** | 15296 (6.7) | 17376 (5.3) | <0.001 |
| **Anxiety disorder** | **Age diagnosis, median [Q1,Q3]** | 50.0 [41.0,60.0] | 53.0 [44.0,62.0] | <0.001 |
|  | **n(%)** | 61069 (26.9) | 159791 (48.8) | <0.001 |
| **Chronic liver disease except cirrhosis** | **Age diagnosis, median [Q1,Q3]** | 56.0 [48.0,65.0] | 60.0 [52.0,69.0] | <0.001 |
|  | **n(%)** | 11604 (5.1) | 11834 (3.6) | <0.001 |
| **Glaucoma** | **Age diagnosis, median [Q1,Q3]** | 63.0 [53.0,71.0] | 63.0 [55.0,71.0] | <0.001 |
|  | **n(%)** | 11356 (5.0) | 21990 (6.7) | <0.001 |
| **Skin melanoma** | **Age diagnosis, median [Q1,Q3]** | 63.0 [52.0,72.0] | 62.0 [53.0,72.0] | 0.369 |
|  | **n(%)** | 1365 (0.6) | 1971 (0.6) | 1.000 |
| **Pure hyperglyceridemia** | **Age diagnosis, median [Q1,Q3]** | 47.0 [40.0,55.0] | 53.0 [46.0,62.0] | <0.001 |
|  | **n(%)** | 42643 (18.8) | 31642 (9.7) | <0.001 |
| **Mixed hyperlipidemia** | **Age diagnosis, median [Q1,Q3]** | 48.0 [41.0,56.0] | 54.0 [47.0,62.0] | <0.001 |
|  | **n(%)** | 75232 (33.1) | 68106 (20.8) | <0.001 |
| **Neurological disease with motor deficit not due to stroke** | **Age diagnosis, median [Q1,Q3]** | 62.0 [53.0,72.0] | 65.0 [54.0,75.0] | <0.001 |
|  | **n(%)** | 13531 (6.0) | 16166 (4.9) | <0.001 |
| **Hypothyroidism** | **Age diagnosis, median [Q1,Q3]** | 55.0 [45.0,66.0] | 55.0 [46.0,64.0] | 0.001 |
|  | **n(%)** | 15738 (6.9) | 86483 (26.4) | <0.001 |
| **Gout and other crystal-induced arthropathies** | **Age diagnosis, median [Q1,Q3]** | 57.0 [48.0,66.0] | 68.0 [58.0,76.0] | <0.001 |
|  | **n(%)** | 23672 (10.4) | 6695 (2.0) | <0.001 |
| **Urinary stones** | **Age diagnosis, median [Q1,Q3]** | 53.0 [44.0,62.0] | 55.0 [46.0,64.0] | <0.001 |
|  | **n(%)** | 35158 (15.5) | 53242 (16.2) | <0.001 |
| **Hepatic steatosis (Fatty liver)** | **Age diagnosis, median [Q1,Q3]** | 55.0 [47.0,63.0] | 59.0 [53.0,67.0] | <0.001 |
|  | **n(%)** | 22779 (10.0) | 25709 (7.8) | <0.001 |
| **Gastroesophageal reflux disease (GERD)** | **Age diagnosis, median [Q1,Q3]** | 54.0 [45.0,64.0] | 59.0 [51.0,67.0] | <0.001 |
|  | **n(%)** | 19867 (8.8) | 38641 (11.8) | <0.001 |
| **Diabetes** | **Age diagnosis, median [Q1,Q3]** | 57.0 [50.0,65.0] | 61.0 [53.0,69.0] | <0.001 |
|  | **n(%)** | 59232 (26.1) | 64433 (19.7) | <0.001 |
| **Collagen disease and vasculitis** | **Age diagnosis, median [Q1,Q3]** | 64.0 [53.0,73.0] | 61.0 [53.0,71.0] | <0.001 |
|  | **n(%)** | 3107 (1.4) | 11820 (3.6) | <0.001 |
| **Ischemic heart disease** | **Age diagnosis, median [Q1,Q3]** | 62.0 [54.0,70.0] | 68.0 [60.0,76.0] | <0.001 |
|  | **n(%)** | 24025 (10.6) | 13811 (4.2) | <0.001 |
| **Mood disorder** | **Age diagnosis, median [Q1,Q3]** | 54.0 [45.0,62.0] | 57.0 [49.0,67.0] | <0.001 |
|  | **n(%)** | 17093 (7.5) | 55672 (17.0) | <0.001 |
| **Retinopathy** | **Age diagnosis, median [Q1,Q3]** | 62.0 [54.0,70.0] | 65.0 [57.0,73.0] | <0.001 |
|  | **n(%)** | 13220 (5.8) | 15093 (4.6) | <0.001 |
| **Occlusion or stenosis of precerebral arteries** | **Age diagnosis, median [Q1,Q3]** | 68.0 [61.0,75.0] | 71.0 [63.0,78.0] | <0.001 |
|  | **n(%)** | 4522 (2.0) | 3080 (0.9) | <0.001 |
| **Tobacco dependence** | **Age diagnosis, median [Q1,Q3]** | 51.0 [43.0,59.0] | 51.0 [44.0,58.0] | <0.001 |
|  | **n(%)** | 51081 (22.5) | 42269 (12.9) | <0.001 |
| **COPD (Chronic Obstructive Pulmonary Disease)** | **Age diagnosis, median [Q1,Q3]** | 62.0 [54.0,69.0] | 62.0 [54.0,70.0] | 0.018 |
|  | **n(%)** | 29918 (13.2) | 26016 (7.9) | <0.001 |
| **Asthma** | **Age diagnosis, median [Q1,Q3]** | 47.0 [37.0,59.0] | 53.0 [43.0,63.0] | <0.001 |
|  | **n(%)** | 17856 (7.9) | 40140 (12.2) | <0.001 |
| **Fibromyalgia** | **Age diagnosis, median [Q1,Q3]** | 52.0 [45.0,58.0] | 53.0 [47.0,59.0] | <0.001 |
|  | **n(%)** | 1381 (0.6) | 30784 (9.4) | <0.001 |
| **Rheumatoid arthritis and related diseases** | **Age diagnosis, median [Q1,Q3]** | 56.0 [47.0,65.0] | 58.0 [50.0,67.0] | <0.001 |
|  | **n(%)** | 3164 (1.4) | 7033 (2.1) | <0.001 |
| **Obesity** | **Age diagnosis, median [Q1,Q3]** | 54.0 [45.0,63.0] | 56.0 [47.0,66.0] | <0.001 |
|  | **n(%)** | 36050 (15.9) | 51941 (15.8) | 0.736 |
| **Acquired valvular disease** | **Age diagnosis, median [Q1,Q3]** | 68.0 [59.0,76.0] | 71.0 [62.0,78.0] | <0.001 |
|  | **n(%)** | 12085 (5.3) | 19046 (5.8) | <0.001 |
| **Atrial fibrillation** | **Age diagnosis, median [Q1,Q3]** | 69.0 [60.0,77.0] | 73.0 [65.0,80.0] | <0.001 |
|  | **n(%)** | 16341 (7.2) | 19301 (5.9) | <0.001 |
| **Malabsorption syndrome and food intolerance** | **Age diagnosis, median [Q1,Q3]** | 54.0 [44.5,63.0] | 56.0 [47.0,65.0] | <0.001 |
|  | **n(%)** | 4603 (2.0) | 12292 (3.8) | <0.001 |
| **Regional enteritis and ulcerative colitis** | **Age diagnosis, median [Q1,Q3]** | 51.0 [41.0,62.0] | 53.0 [43.0,64.0] | <0.001 |
|  | **n(%)** | 2230 (1.0) | 2614 (0.8) | <0.001 |
| **Other arthropathy** | **Age diagnosis, median [Q1,Q3]** | 57.0 [48.0,65.0] | 59.0 [51.0,68.0] | <0.001 |
|  | **n(%)** | 18585 (8.2) | 29069 (8.9) | <0.001 |
| **Atopic dermatitis** | **Age diagnosis, median [Q1,Q3]** | 52.0 [41.0,63.0] | 53.0 [43.0,63.0] | 0.038 |
|  | **n(%)** | 2003 (0.9) | 4316 (1.3) | <0.001 |
| **Alcohol dependence** | **Age diagnosis, median [Q1,Q3]** | 56.0 [47.0,65.0] | 55.0 [46.0,63.0] | <0.001 |
|  | **n(%)** | 23040 (10.2) | 5024 (1.5) | <0.001 |
| **Intellectual disability** | **Age diagnosis, median [Q1,Q3]** | 43.0 [33.0,52.0] | 45.0 [37.0,54.0] | <0.001 |
|  | **n(%)** | 991 (0.4) | 1111 (0.3) | <0.001 |
| **Childhood and adolescence-onset disorder** | **Age diagnosis, median [Q1,Q3]** | 56.0 [44.0,69.0] | 63.0 [51.0,75.0] | <0.001 |
|  | **n(%)** | 2153 (0.9) | 3265 (1.0) | 0.078 |
| **Other functional disorder** | **Age diagnosis, median [Q1,Q3]** | 52.0 [43.0,62.0] | 57.0 [49.0,68.0] | <0.001 |
|  | **n(%)** | 2106 (0.9) | 3069 (0.9) | 0.754 |
| **Other organic mental disorder** | **Age diagnosis, median [Q1,Q3]** | 70.0 [57.0,79.0] | 76.0 [68.0,82.0] | <0.001 |
|  | **n(%)** | 9966 (4.4) | 16686 (5.1) | <0.001 |
| **Liver cirrhosis** | **Age diagnosis, median [Q1,Q3]** | 59.0 [52.0,67.0] | 61.0 [53.0,70.0] | <0.001 |
|  | **n(%)** | 1748 (0.8) | 1203 (0.4) | <0.001 |
| **Peripheral arterial disease** | **Age diagnosis, median [Q1,Q3]** | 65.0 [57.0,73.0] | 67.0 [57.0,76.0] | <0.001 |
|  | **n(%)** | 17094 (7.5) | 15641 (4.8) | <0.001 |
| **Colorectal cancer** | **Age diagnosis, median [Q1,Q3]** | 66.0 [59.0,73.0] | 66.0 [58.0,74.0] | 0.681 |
|  | **n(%)** | 5126 (2.3) | 5038 (1.5) | <0.001 |
| **Heart failure** | **Age diagnosis, median [Q1,Q3]** | 70.0 [61.0,78.0] | 75.0 [67.0,82.0] | <0.001 |
|  | **n(%)** | 17550 (7.7) | 23626 (7.2) | <0.001 |
| **Dementia** | **Age diagnosis, median [Q1,Q3]** | 77.0 [70.0,83.0] | 78.0 [72.0,84.0] | <0.001 |
|  | **n(%)** | 5165 (2.3) | 13286 (4.1) | <0.001 |
| **Dependence on other substances** | **Age diagnosis, median [Q1,Q3]** | 44.0 [36.0,53.0] | 55.0 [45.0,66.0] | <0.001 |
|  | **n(%)** | 3797 (1.7) | 2101 (0.6) | <0.001 |
| **Congenital cardiac and circulatory anomaly** | **Age diagnosis, median [Q1,Q3]** | 57.0 [48.0,66.0] | 59.0 [47.0,70.0] | <0.001 |
|  | **n(%)** | 1484 (0.7) | 1552 (0.5) | <0.001 |
| **Aneurysm of aorta, peripheral and visceral arteries** | **Age diagnosis, median [Q1,Q3]** | 67.0 [59.0,74.0] | 69.0 [61.0,76.0] | <0.001 |
|  | **n(%)** | 4684 (2.1) | 2382 (0.7) | <0.001 |
| **Kidney and renal pelvis cancer** | **Age diagnosis, median [Q1,Q3]** | 64.0 [56.0,72.8] | 67.0 [58.0,74.0] | <0.001 |
|  | **n(%)** | 1202 (0.5) | 895 (0.3) | <0.001 |
| **Intra-abdominal arteriopathy** | **Age diagnosis, median [Q1,Q3]** | 69.5 [60.0,77.0] | 73.0 [64.0,81.0] | <0.001 |
|  | **n(%)** | 650 (0.3) | 998 (0.3) | 0.232 |
| **Schizophrenic disorder** | **Age diagnosis, median [Q1,Q3]** | 44.0 [35.0,54.0] | 53.0 [43.0,66.0] | <0.001 |
|  | **n(%)** | 3529 (1.6) | 3727 (1.1) | <0.001 |
| **Bladder cancer** | **Age diagnosis, median [Q1,Q3]** | 67.0 [60.0,74.0] | 66.0 [58.0,74.0] | 0.001 |
|  | **n(%)** | 4772 (2.1) | 1451 (0.4) | <0.001 |
| **Parkinson's disease** | **Age diagnosis, median [Q1,Q3]** | 72.0 [64.0,78.0] | 74.0 [67.0,79.0] | <0.001 |
|  | **n(%)** | 2310 (1.0) | 3468 (1.1) | 0.148 |
| **Epilepsy** | **Age diagnosis, median [Q1,Q3]** | 54.0 [42.0,66.0] | 57.0 [44.0,70.0] | <0.001 |
|  | **n(%)** | 4531 (2.0) | 5816 (1.8) | <0.001 |
| **Non-Hodgkin lymphoma** | **Age diagnosis, median [Q1,Q3]** | 62.0 [51.0,71.0] | 62.0 [54.0,71.0] | 0.038 |
|  | **n(%)** | 1171 (0.5) | 1404 (0.4) | <0.001 |
| **Age-related macular degeneration** | **Age diagnosis, median [Q1,Q3]** | 73.0 [63.5,79.0] | 75.0 [67.0,81.0] | <0.001 |
|  | **n(%)** | 3983 (1.8) | 8390 (2.6) | <0.001 |
| **Other developmental disorder** | **Age diagnosis, median [Q1,Q3]** | 58.0 [45.0,69.0] | 61.0 [51.0,73.2] | <0.001 |
|  | **n(%)** | 518 (0.2) | 664 (0.2) | 0.045 |
| **Poorly defined and other cerebrovascular diseases** | **Age diagnosis, median [Q1,Q3]** | 69.0 [60.0,77.0] | 72.0 [62.0,79.0] | <0.001 |
|  | **n(%)** | 3278 (1.4) | 3702 (1.1) | <0.001 |
| **Pancreatic cancer** | **Age diagnosis, median [Q1,Q3]** | 68.0 [59.2,75.0] | 70.0 [62.2,78.0] | 0.001 |
|  | **n(%)** | 382 (0.2) | 478 (0.1) | 0.040 |
| **Bronchus and lung cancer** | **Age diagnosis, median [Q1,Q3]** | 69.0 [62.0,76.0] | 65.0 [59.0,73.0] | <0.001 |
|  | **n(%)** | 2335 (1.0) | 1129 (0.3) | <0.001 |
| **Adult personality and behavioral disorder** | **Age diagnosis, median [Q1,Q3]** | 48.0 [39.0,58.0] | 53.0 [45.0,63.0] | <0.001 |
|  | **n(%)** | 3949 (1.7) | 5591 (1.7) | 0.346 |
| **Thyroid cancer** | **Age diagnosis, median [Q1,Q3]** | 55.0 [45.0,63.0] | 55.0 [47.0,64.0] | 0.398 |
|  | **n(%)** | 423 (0.2) | 1600 (0.5) | <0.001 |
| **Leukemia** | **Age diagnosis, median [Q1,Q3]** | 64.0 [54.0,74.0] | 66.0 [58.0,75.0] | 0.001 |
|  | **n(%)** | 759 (0.3) | 768 (0.2) | <0.001 |
| **Eating behavior disorder** | **Age diagnosis, median [Q1,Q3]** | 51.5 [40.0,69.0] | 47.0 [37.0,56.0] | <0.001 |
|  | **n(%)** | 240 (0.1) | 1469 (0.4) | <0.001 |
| **Stomach cancer** | **Age diagnosis, median [Q1,Q3]** | 68.0 [59.0,76.0] | 70.0 [61.0,77.0] | 0.039 |
|  | **n(%)** | 463 (0.2) | 404 (0.1) | <0.001 |
| **Immunoproliferative cancer** | **Age diagnosis, median [Q1,Q3]** | 67.0 [59.0,75.0] | 68.0 [60.0,75.0] | 0.589 |
|  | **n(%)** | 348 (0.2) | 404 (0.1) | 0.003 |
| **HIV** | **Age diagnosis, median [Q1,Q3]** | 42.0 [36.0,49.0] | 41.0 [35.0,49.0] | 0.055 |
|  | **n(%)** | 1399 (0.6) | 590 (0.2) | <0.001 |
| **Head and neck cancer** | **Age diagnosis, median [Q1,Q3]** | 63.0 [55.0,70.0] | 63.0 [55.0,72.0] | 0.172 |
|  | **n(%)** | 2213 (1.0) | 1240 (0.4) | <0.001 |
| **Sequelae of cerebrovascular disease** | **Age diagnosis, median [Q1,Q3]** | 68.0 [60.0,76.0] | 74.0 [64.0,81.8] | <0.001 |
|  | **n(%)** | 2804 (1.2) | 2470 (0.8) | <0.001 |
| **Bone and soft tissue cancer** | **Age diagnosis, median [Q1,Q3]** | 61.0 [51.0,71.0] | 62.0 [53.0,70.0] | 0.750 |
|  | **n(%)** | 418 (0.2) | 537 (0.2) | 0.078 |
| **Autism spectrum disorder** | **Age diagnosis, median [Q1,Q3]** | 42.0 [29.5,53.0] | 53.0 [45.0,62.0] | <0.001 |
|  | **n(%)** | 104 (0.0) | 65 (0.0) | <0.001 |
| **Hodgkin's disease** | **Age diagnosis, median [Q1,Q3]** | 49.0 [35.5,60.0] | 48.5 [37.0,60.8] | 0.736 |
|  | **n(%)** | 251 (0.1) | 246 (0.1) | <0.001 |
| **Liver and biliary tract cancer** | **Age diagnosis, median [Q1,Q3]** | 67.0 [59.0,75.0] | 71.0 [63.0,78.0] | <0.001 |
|  | **n(%)** | 556 (0.2) | 380 (0.1) | <0.001 |
| **Kaposi's sarcoma** | **Age diagnosis, median [Q1,Q3]** | 64.0 [47.0,73.0] | 70.0 [56.0,82.5] | 0.126 |
|  | **n(%)** | 49 (0.0) | 19 (0.0) | <0.001 |
