## Supplementary Table 2 for "The Impact of Menopause on Hypercholesterolemia and Comorbidities: A Population-Based Study"

**Supplementary Table 2**. Comorbidity categories used and diseases included in them.

| **Comorbidity category** | **Disease** |
| --- | --- |
| Cancers | - Skin melanoma, - Bladder cancer - Colorectal cancer, - Kidney and renal pelvis cancer, - Pancreatic cancer, - Bronchus and lung cancer, - Thyroid cancer, - Leukemia, - Stomach cancer, - Immunoproliferative cancer, - Head and neck cancer, - Bone and soft tissue cancer, - Liver and biliary tract cancer, - Kaposi's sarcoma, - Hodgkin's disease, - Non-Hodgkin lymphoma |
| Cardiovascular Diseases | - Hypertension, - Ischemic heart disease, - Acquired valvular disease, - Atrial fibrillation, - Heart failure, - Peripheral arterial disease, - Aneurysm of aorta, peripheral and visceral arteries, - Intra-abdominal arteriopathy |
| Endocrine and Metabolic Disorders: | - Diabetes, - Hypothyroidism, - Pure hyperglyceridemia, - Mixed hyperlipidemia, - Obesity, - Gout and other crystal-induced arthropathies |
| Bone and Joint Disorders: | - Osteoporosis, - Osteoarthritis, spondylosis, - Rheumatoid arthritis and related diseases, - Other arthropathy, - Fibromyalgia |
| Neurological Disorders: | - Extrapyramidal syndrome, - Neurological disease with motor deficit not due to stroke, - Parkinson's disease, - Epilepsy, - Sequelae of cerebrovascular disease, - Dementia |
| Respiratory Diseases: | - COPD (Chronic Obstructive Pulmonary Disease), - Asthma |
| Mental Health and Substance Use: | - Anxiety disorder, - Mood disorder, - Schizophrenic disorder, - Adult personality and behavioral disorder, - Other organic mental disorder, - Other functional disorder, - Intellectual disability, - Childhood and adolescence-onset disorder, - Dependence on other substances, - Alcohol dependence, - Tobacco dependence, - Autism spectrum disorder, - Other developmental disorder, - Eating behavior disorder |
| Infectious Diseases: | - HIV |
| Liver Diseases: | - Chronic liver disease except cirrhosis - Hepatic steatosis (Fatty liver), - Liver cirrhosis |
| Gastrointestinal Disorders: | - Gastroesophageal reflux disease (GERD), - Malabsorption syndrome and food intolerance, - Regional enteritis and ulcerative colitis |
| Ocular Diseases: | - Glaucoma, - Retinopathy, - Age-related macular degeneration |
| Renal and Urinary Disorders: | - Chronic renal insufficiency, - Urinary stones |
| Dermatological Disorders: | - Psoriasis, - Atopic dermatitis |
| Cerebrovascular Disorders: | - Occlusion or stenosis of precerebral arteries, - Poorly defined and other cerebrovascular diseases |
| Congenital and Developmental Disorders: | - Congenital cardiac and circulatory anomaly |
