## Supplementary Table 3 for "The Impact of Menopause on Hypercholesterolemia and Comorbidities: A Population-Based Study"

**Supplementary Table 3**. Sex-independent and sex-dependent comorbidities.

| **Sex-independent** | **Sex-dependent** |
| --- | --- |
| - Acquired valvular disease - Alcohol dependence - Aneurysm of aorta, peripheral and visceral arteries - Autism spectrum disorder - Bladder cancer - Bone and soft tissue cancer - Bronchus and lung cancer - COPD (Chronic Obstructive Pulmonary Disease) - Colorectal cancer - Congenital cardiac and circulatory anomaly - Eating behavior disorder - Epilepsy - Fibromyalgia - HIV - Head and neck cancer - Hodgkin's disease - Immunoproliferative cancer - Intellectual disability - Kaposi's sarcoma - Kidney and renal pelvis cancer - Liver and biliary tract cancer - Liver cirrhosis - Neurological disease with motor deficit not due to stroke - Occlusion or stenosis of precerebral arteries - Pancreatic cancer - Peripheral arterial disease - Poorly defined and other cerebrovascular diseases - Sequelae of cerebrovascular disease - Stomach cancer - Thyroid cancer | - Adult personality and behavioral disorder - Age-related macular degeneration - Anxiety disorder - Asthma - Atopic dermatitis - Atrial fibrillation - Childhood and adolescence-onset disorder - Chronic liver disease except cirrhosis - Chronic renal insufficiency - Collagen disease and vasculitis - Dementia - Dependence on other substances - Diabetes - Extrapyramidal syndrome - Gastroesophageal reflux disease (GERD) - Glaucoma - Gout and other crystal-induced arthropathies - Heart failure - Hepatic steatosis (Fatty liver) - Hypertension - Hypothyroidism - Intra-abdominal arteriopathy - Ischemic heart disease - Leukemia - Malabsorption syndrome and food intolerance - Mixed hyperlipidemia - Mood disorder - Non-Hodgkin lymphoma - Obesity - Osteoarthritis, spondylosis - Osteoporosis - Other arthropathy - Other developmental disorder - Other functional disorder - Other organic mental disorder - Parkinson's disease - Psoriasis - Pure hyperglyceridemia - Regional enteritis and ulcerative colitis - Retinopathy - Rheumatoid arthritis and related diseases - Schizophrenic disorder - Skin melanoma - Tobacco dependence - Urinary stones |
