## Supplementary Table 4 for "The Impact of Menopause on Hypercholesterolemia and Comorbidities: A Population-Based Study"

**Supplementary Table 4**. **Comorbidities in woman**. Number of patients with at least one comorbidity (second and third columns represent observations in patients younger than 50 years, and fourth and fifth columns in patients 50 years old and older). A X^2^ test has been conducted to detect significant increases of comorbidity diagnoses in women with respect to men when the older (post-menopausal age) are compared to the younger, and the sixth column reports the p-value. The seventh column represent the ratio woman/man between the respective increase in comorbidities in older with respect to younger. The last column is the percentage of woman with the comorbidity with respect to all women with comorbidities.

|  | **<50** |  | **≥50** |  |  |  |
| --- | --- | --- | --- | --- | --- | --- |
| **Comorbidities** | **Men** | **Women** | **Men** | **Women** | **X2 p-Value** | **Women/men** |
| Acquired valvular disease | 1,057 | 1,436 | 11,028 | 17,610 | <0.001 | 1.18 |
| Adult personality and behavioral disorder | 2,080 | 2,153 | 1,869 | 3,438 | <0.001 | 1.78 |
| Age-related macular degeneration | 216 | 232 | 3,767 | 8,158 | <0.001 | 2.02 |
| Alcohol dependence | 6,952 | 1,671 | 16,088 | 3,353 | <0.001 | 0.87 |
| Aneurysm of aorta, peripheral and visceral arteries | 359 | 184 | 4,325 | 2,198 | 0.966 | 0.99 |
| Anxiety disorder | 29,695 | 64,875 | 31,374 | 94,916 | <0.001 | 1.38 |
| Asthma | 9,922 | 16,714 | 7,934 | 23,426 | <0.001 | 1.75 |
| Atopic dermatitis | 874 | 1,743 | 1,129 | 2,573 | 0.016 | 1.14 |
| Atrial fibrillation | 1,268 | 514 | 15,073 | 18,787 | <0.001 | 3.07 |
| Autism spectrum disorder | 71 | 25 | 33 | 40 | <0.001 | 3.44 |
| Bladder cancer | 261 | 102 | 4,511 | 1,349 | 0.031 | 0.77 |
| Bone and soft tissue cancer | 89 | 91 | 329 | 446 | 0.105 | 1.33 |
| Bronchus and lung cancer | 81 | 56 | 2,254 | 1,073 | 0.044 | 0.69 |
| Childhood and adolescence-onset disorder | 763 | 712 | 1,390 | 2,553 | <0.001 | 1.97 |
| Chronic liver disease except cirrhosis | 3,488 | 2,127 | 8,116 | 9,707 | <0.001 | 1.96 |
| Chronic renal insufficiency | 1,233 | 745 | 14,063 | 16,631 | <0.001 | 1.96 |
| Collagen disease and vasculitis | 588 | 2,108 | 2,519 | 9,712 | 0.168 | 1.08 |
| Colorectal cancer | 344 | 393 | 4,782 | 4,645 | 0.038 | 0.85 |
| Congenital cardiac and circulatory anomaly | 433 | 442 | 1,051 | 1,110 | 0.7 | 1.03 |
| COPD (Chronic Obstructive Pulmonary Disease) | 4,426 | 3,929 | 25,492 | 22,087 | 0.313 | 0.98 |
| Dementia | 107 | 118 | 5,058 | 13,168 | <0.001 | 2.36 |
| Dependence on other substances | 2,502 | 758 | 1,295 | 1,343 | <0.001 | 3.42 |
| Diabetes | 14,358 | 10,845 | 44,874 | 53,588 | <0.001 | 1.58 |
| Eating behavior disorder | 113 | 860 | 127 | 609 | 0.001 | 0.63 |
| Epilepsy | 1,829 | 2,034 | 2,702 | 3,782 | <0.001 | 1.26 |
| Extrapyramidal syndrome | 602 | 681 | 3,714 | 7,407 | <0.001 | 1.76 |
| Fibromyalgia | 558 | 10,749 | 823 | 20,035 | <0.001 | 1.26 |
| Gastroesophageal reflux disease (GERD) | 7,384 | 7,999 | 12,483 | 30,642 | <0.001 | 2.27 |
| Glaucoma | 1,932 | 2,861 | 9,424 | 19,129 | <0.001 | 1.37 |
| Gout and other crystal-induced arthropathies | 6,607 | 515 | 17,065 | 6,180 | <0.001 | 4.65 |
| Head and neck cancer | 270 | 174 | 1,943 | 1,066 | 0.136 | 0.85 |
| Heart failure | 1,138 | 711 | 16,412 | 22,915 | <0.001 | 2.23 |
| Hepatic steatosis (Fatty liver) | 7,472 | 4,014 | 15,307 | 21,695 | <0.001 | 2.64 |
| HIV | 1,050 | 448 | 349 | 142 | 0.72 | 0.95 |
| Hodgkin's disease | 131 | 129 | 120 | 117 | 1 | 0.99 |
| Hypertension | 39,138 | 38,776 | 91,532 | 147,538 | <0.001 | 1.63 |
| Hypothyroidism | 5,577 | 28,855 | 10,161 | 57,628 | <0.001 | 1.10 |
| Immunoproliferative cancer | 30 | 24 | 318 | 380 | 0.201 | 1.49 |
| Intellectual disability | 680 | 696 | 311 | 415 | 0.005 | 1.30 |
| Intra-abdominal arteriopathy | 57 | 49 | 593 | 949 | 0.003 | 1.86 |
| Ischemic heart disease | 3,242 | 789 | 20,783 | 13,022 | <0.001 | 2.57 |
| Kaposi's sarcoma | 13 | 4 | 36 | 15 | 0.761 | 1.35 |
| Kidney and renal pelvis cancer | 140 | 89 | 1,062 | 806 | 0.244 | 1.19 |
| Leukemia | 135 | 94 | 624 | 674 | 0.003 | 1.55 |
| Liver and biliary tract cancer | 22 | 15 | 534 | 365 | 1 | 1.00 |
| Liver cirrhosis | 314 | 188 | 1,434 | 1,015 | 0.107 | 1.18 |
| Malabsorption syndrome and food intolerance | 1,741 | 3,840 | 2,862 | 8,452 | <0.001 | 1.34 |
| Metabolic syndrome | 2,488 | 2,073 | 11,953 | 15,690 | <0.001 | 1.58 |
| Mixed hyperlipidemia | 41,025 | 22,744 | 34,207 | 45,362 | <0.001 | 2.39 |
| Mood disorder | 6,295 | 14,456 | 10,798 | 41,216 | <0.001 | 1.66 |
| Neurological disease with motor deficit not due to stroke | 2,486 | 2,751 | 11,045 | 13,415 | 0.002 | 1.10 |
| Non-Hodgkin lymphoma | 250 | 225 | 921 | 1,179 | 0.001 | 1.42 |
| Obesity | 13,278 | 15,701 | 22,772 | 36,240 | <0.001 | 1.35 |
| Occlusion or stenosis of precerebral arteries | 149 | 119 | 4,373 | 2,961 | 0.209 | 0.85 |
| Osteoarthritis, spondylosis | 14,247 | 28,405 | 66,032 | 156,067 | <0.001 | 1.19 |
| Osteoporosis | 322 | 2,081 | 2,623 | 42,255 | <0.001 | 2.49 |
| Other arthropathy | 5,452 | 5,943 | 13,133 | 23,126 | <0.001 | 1.62 |
| Other developmental disorder | 169 | 151 | 349 | 513 | <0.001 | 1.65 |
| Other functional disorder | 916 | 829 | 1,190 | 2,240 | <0.001 | 2.08 |
| Other organic mental disorder | 1,435 | 670 | 8,531 | 16,016 | <0.001 | 4.02 |
| Pancreatic cancer | 27 | 20 | 355 | 458 | 0.09 | 1.74 |
| Parkinson's disease | 88 | 59 | 2,222 | 3,409 | <0.001 | 2.29 |
| Peripheral arterial disease | 1,518 | 1,852 | 15,576 | 13,789 | <0.001 | 0.73 |
| Poorly defined and other cerebrovascular diseases | 197 | 235 | 3,081 | 3,467 | 0.592 | 0.94 |
| Psoriasis | 4,602 | 4,524 | 6,049 | 7,921 | <0.001 | 1.33 |
| Pure hyperglyceridemia | 24,515 | 11,339 | 18,128 | 20,303 | <0.001 | 2.42 |
| Regional enteritis and ulcerative colitis | 1,034 | 1,058 | 1,196 | 1,556 | <0.001 | 1.27 |
| Retinopathy | 2,150 | 1,535 | 11,070 | 13,558 | <0.001 | 1.72 |
| Rheumatoid arthritis and related diseases | 946 | 1,612 | 2,218 | 5,421 | <0.001 | 1.43 |
| Schizophrenic disorder | 2,294 | 1,510 | 1,235 | 2,217 | <0.001 | 2.73 |
| Sequelae of cerebrovascular disease | 197 | 127 | 2,607 | 2,343 | 0.005 | 1.39 |
| Skin melanoma | 272 | 359 | 1,093 | 1,612 | 0.231 | 1.12 |
| Stomach cancer | 29 | 23 | 434 | 381 | 0.834 | 1.11 |
| Thyroid cancer | 135 | 513 | 288 | 1,087 | 1 | 0.99 |
| Tobacco dependence | 22,162 | 18,437 | 28,919 | 23,832 | 0.48 | 0.99 |
| Urinary stones | 14,128 | 18,296 | 21,030 | 34,946 | <0.001 | 1.28 |
