## Supplementary Figure 1 for "The Impact of Menopause on Hypercholesterolemia and Comorbidities: A Population-Based Study"

| **A**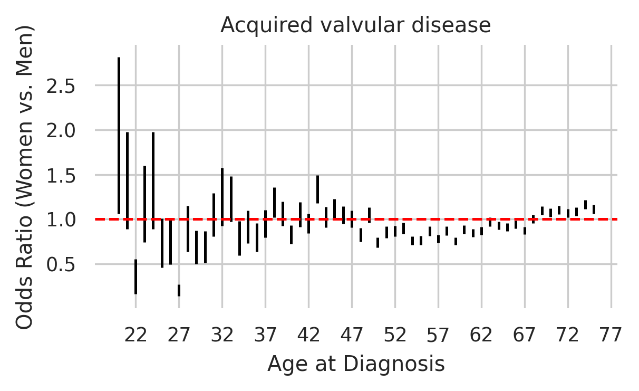 | **B**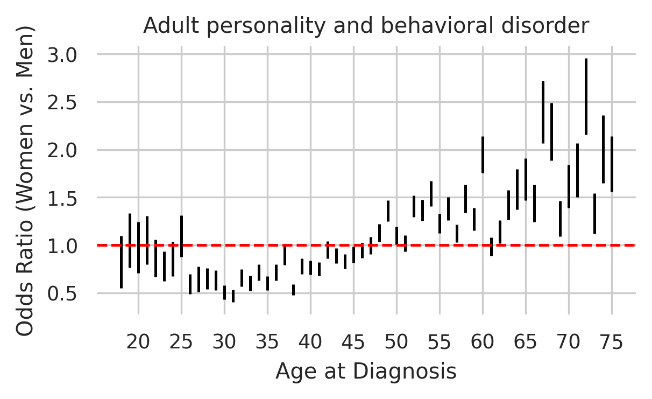 |
| --- | --- |
| **C**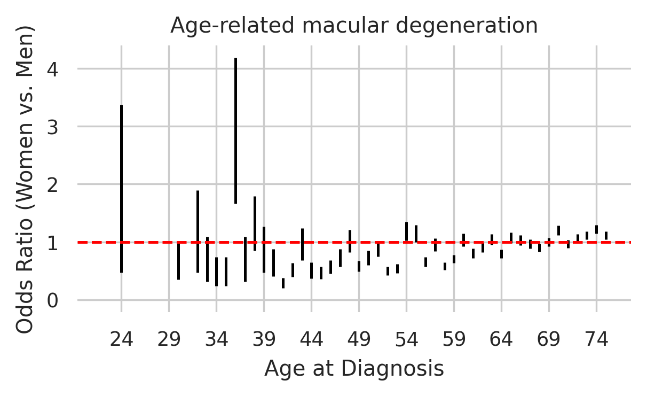 | **D**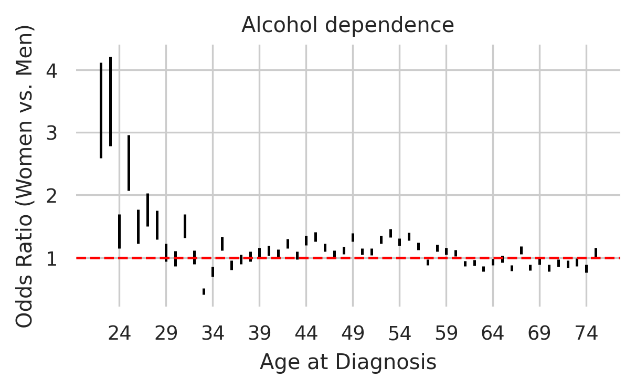 |
| **E**  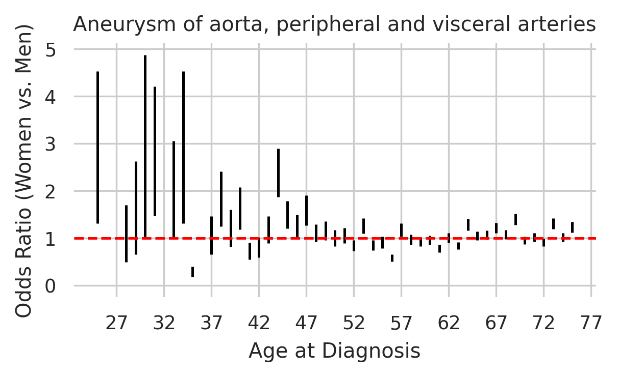 | **F**  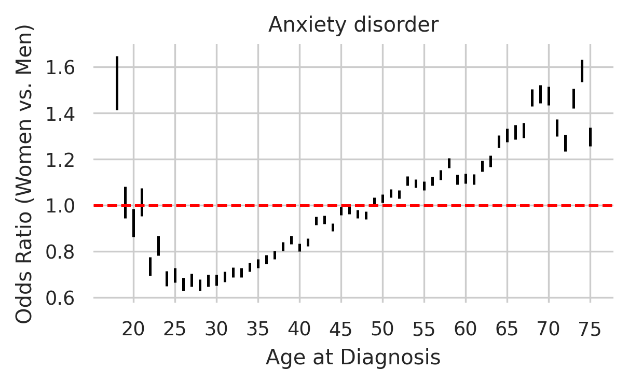 |
| **G**  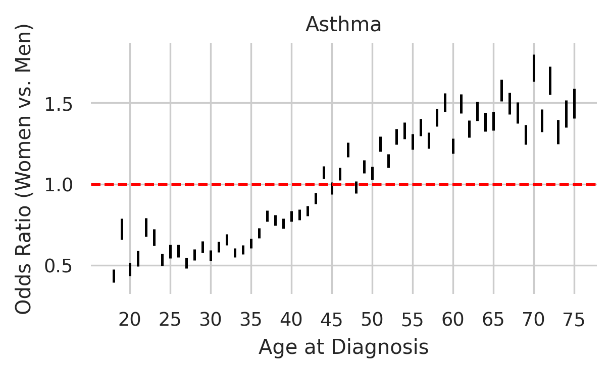 | H  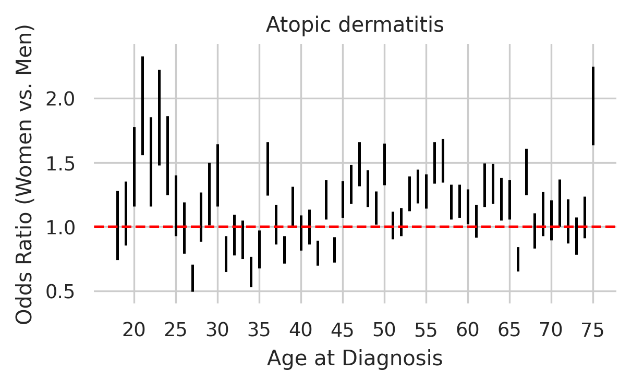 |
| **I**  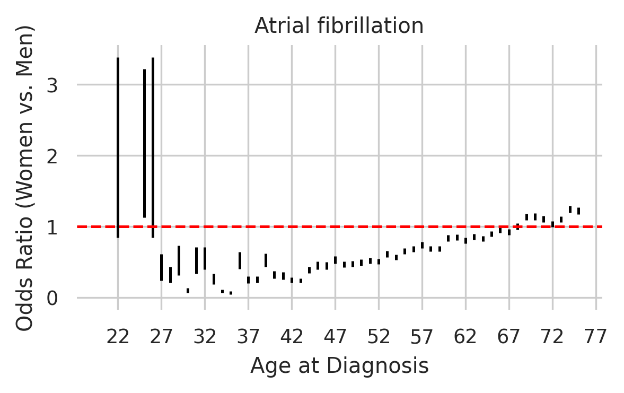 | **J**  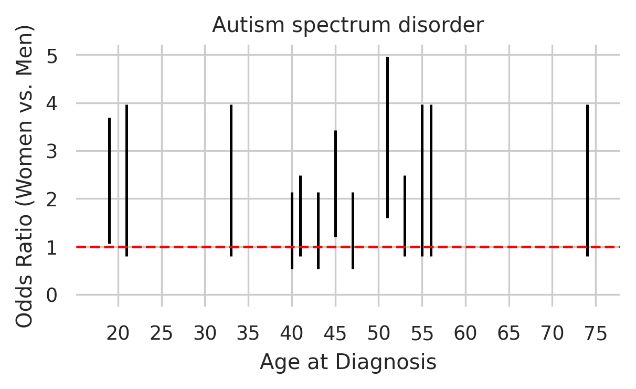 |
| **K**  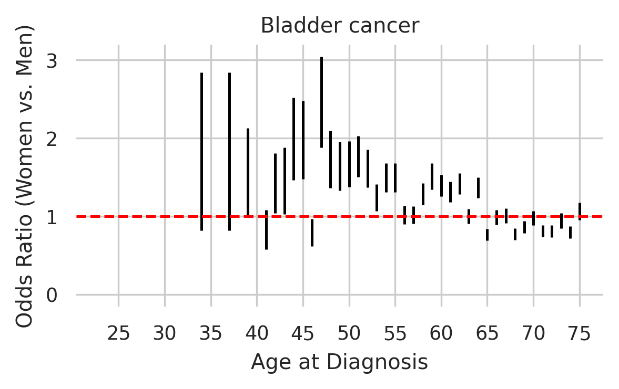 | **L**  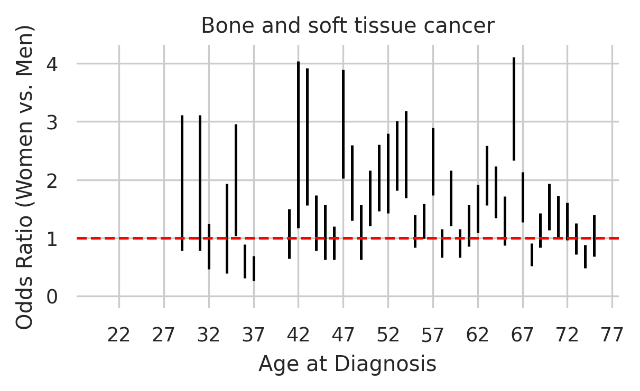 |
| **M**  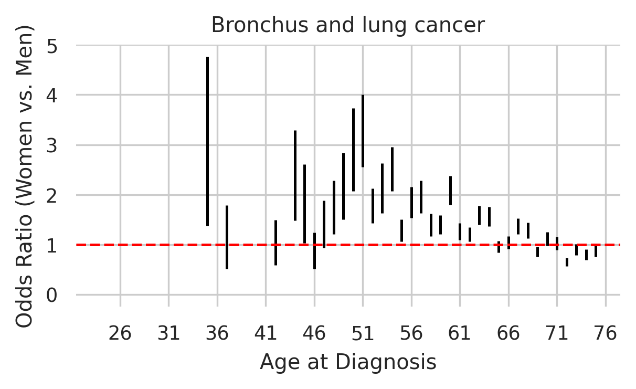 | **N**  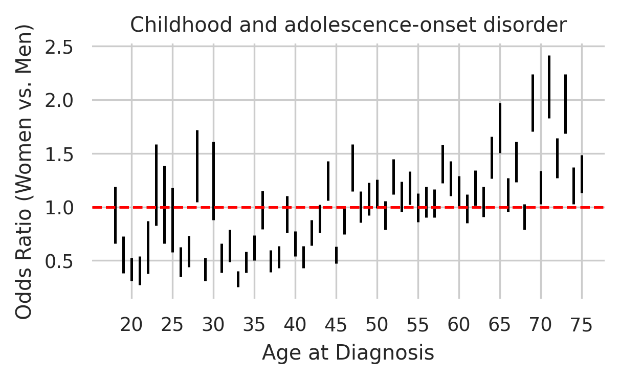 |
| **O**  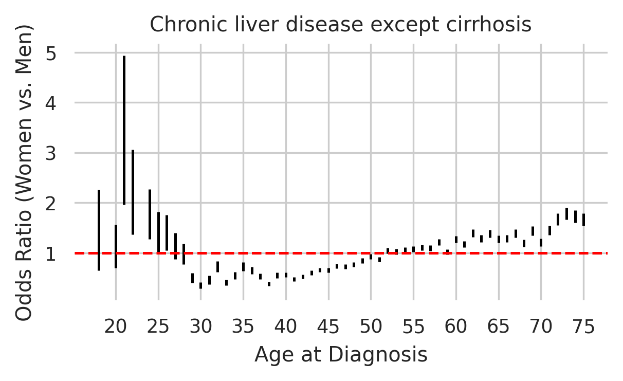 | **P**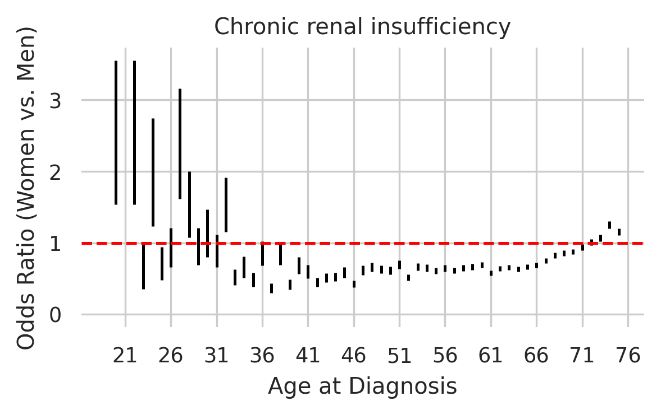 |
| Q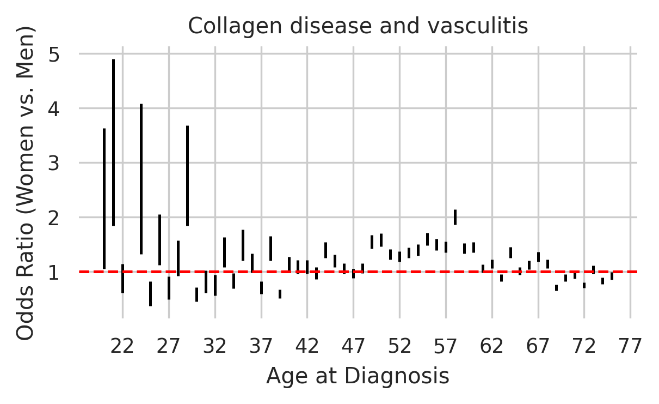 | R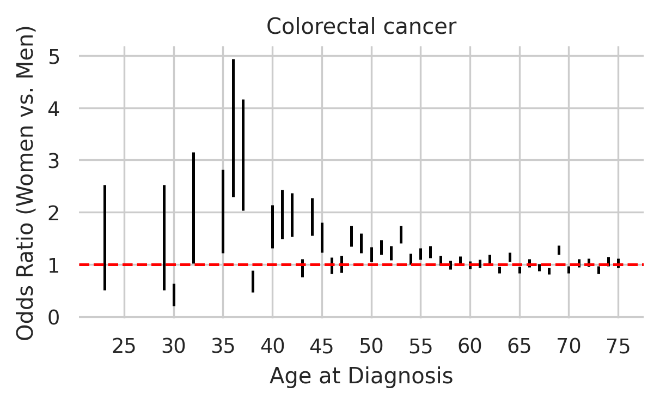 |
| **S**  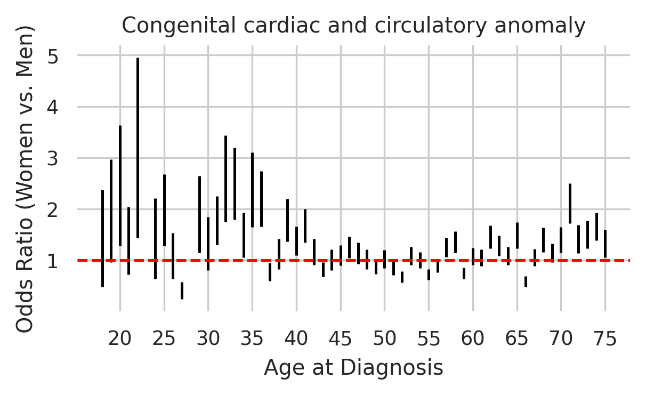 | **T**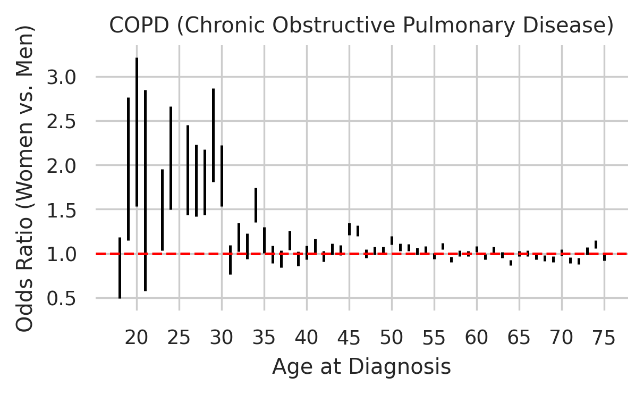 |
| **U**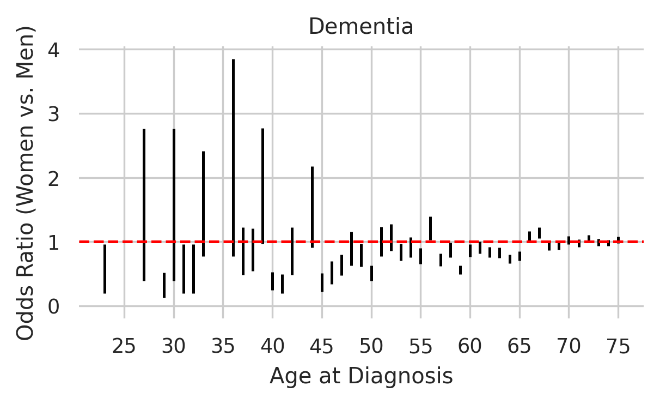 | **V**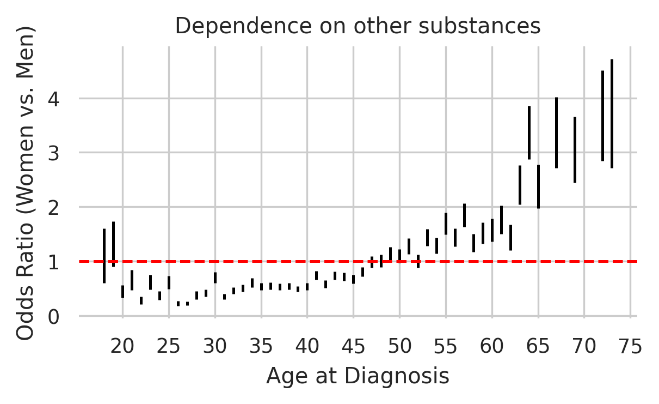 |
| W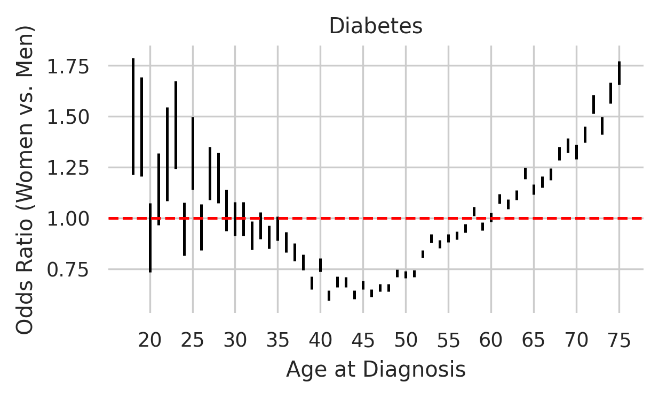 | X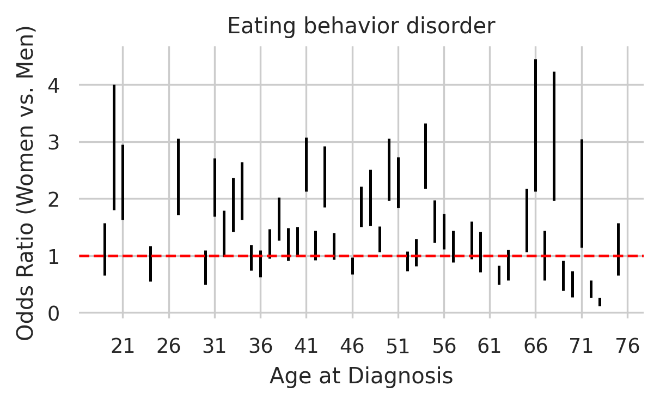 |
| **Y**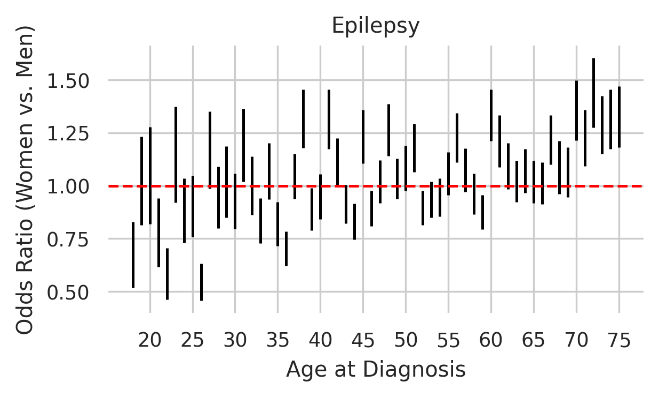 | **Z**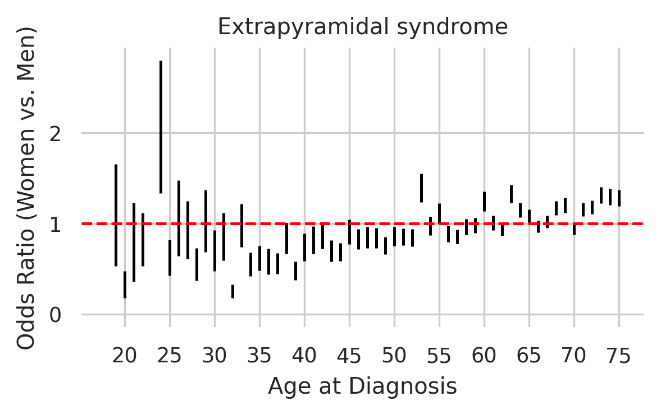 |
| **AA**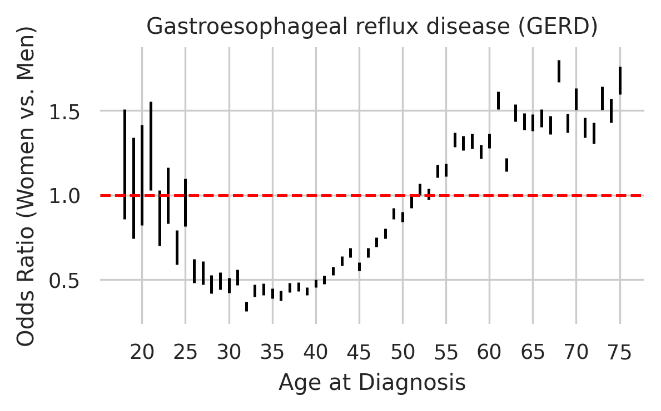 | **AB**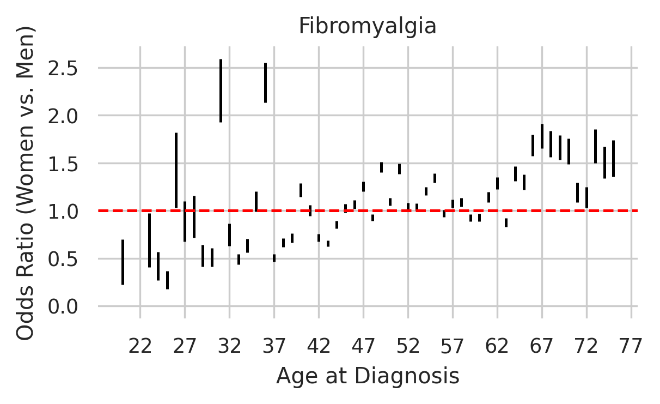 |
| **AC**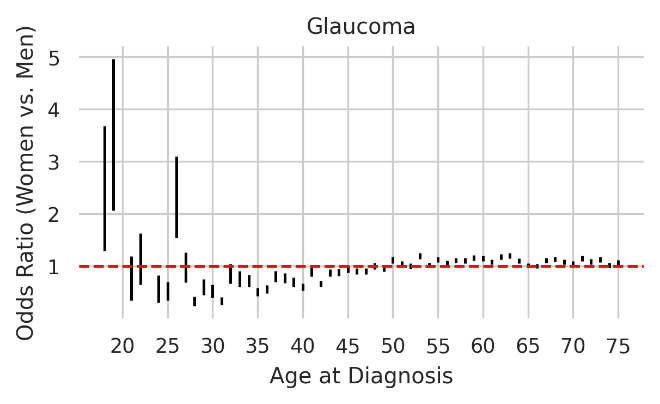 | **AD**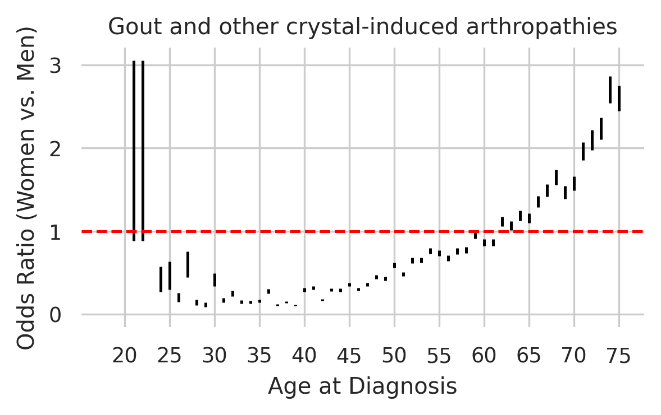 |
| **AF**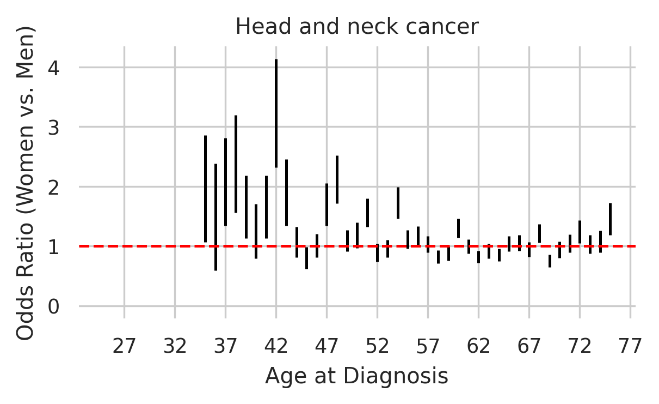 | **AG**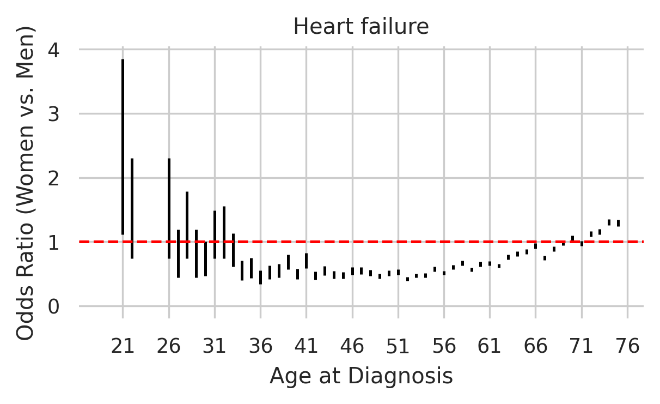 |
| **AH**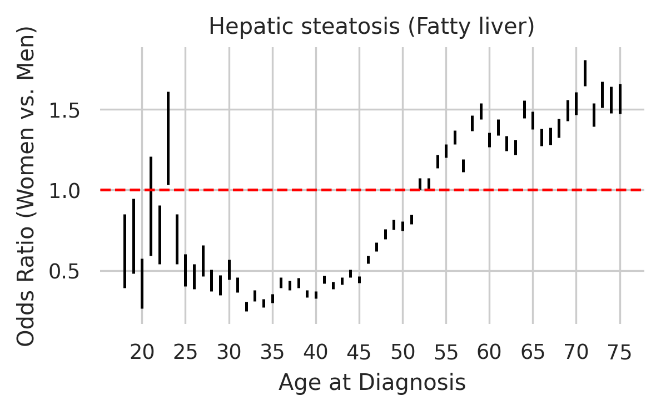 | **AI**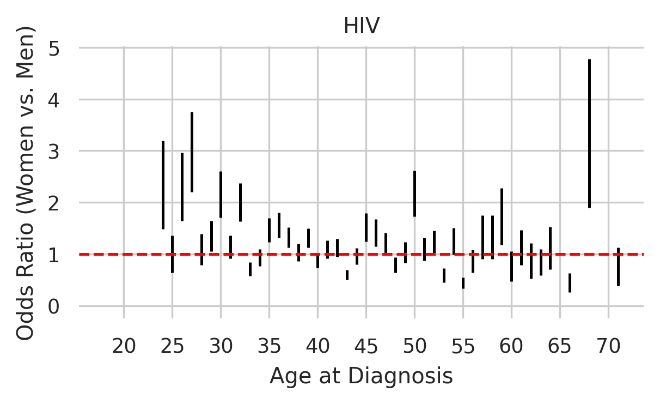 |
| **AJ**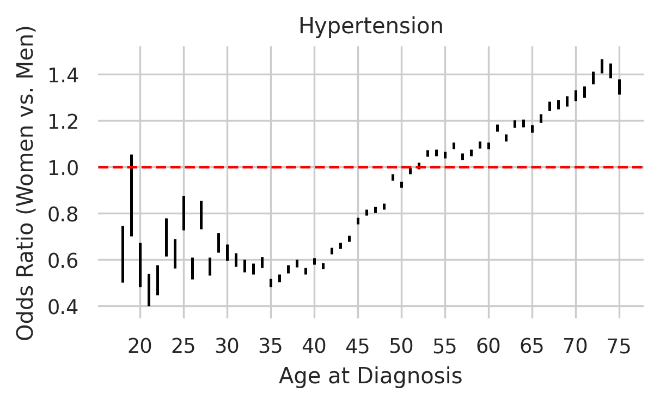 | **AK**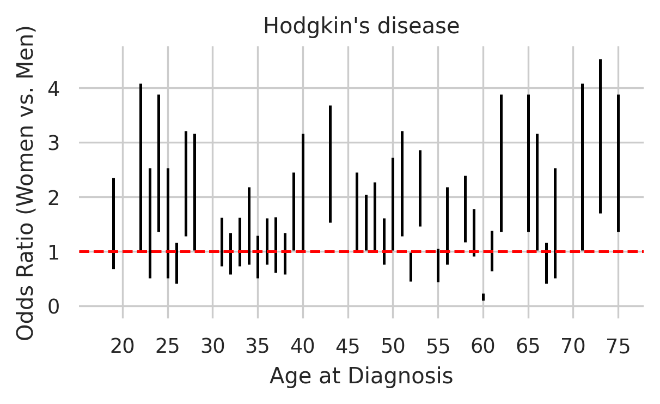 |
| **AL**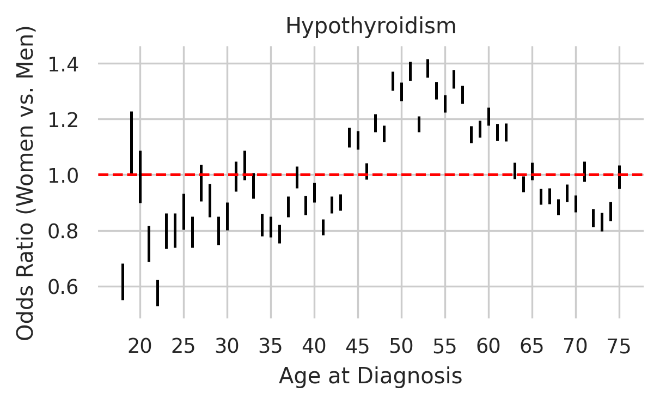 | **AM**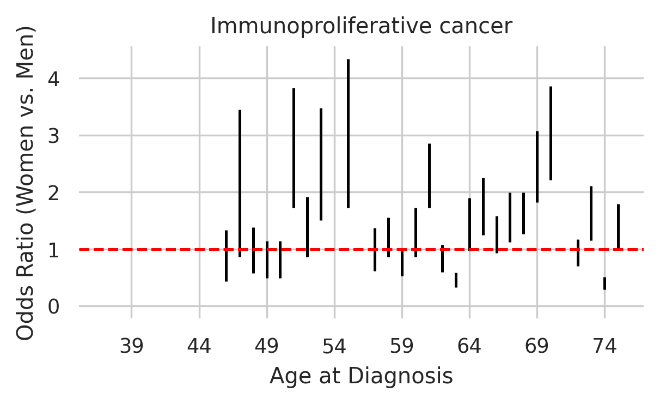 |
| **AN**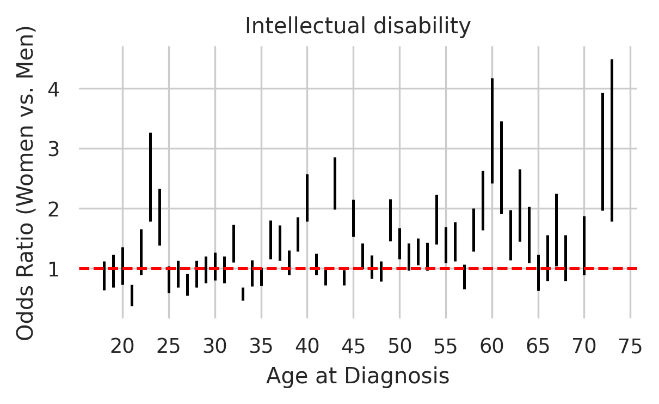**AP**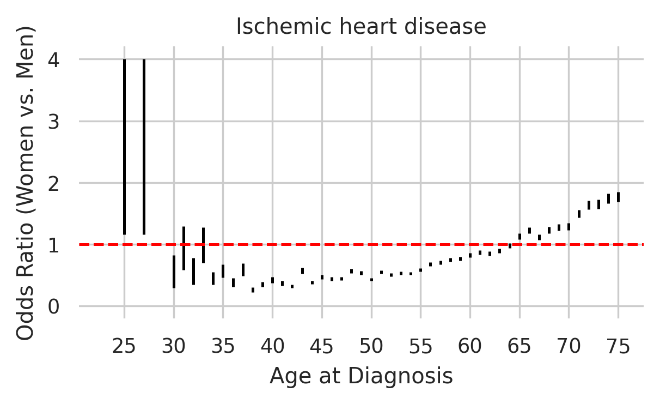 | **AO**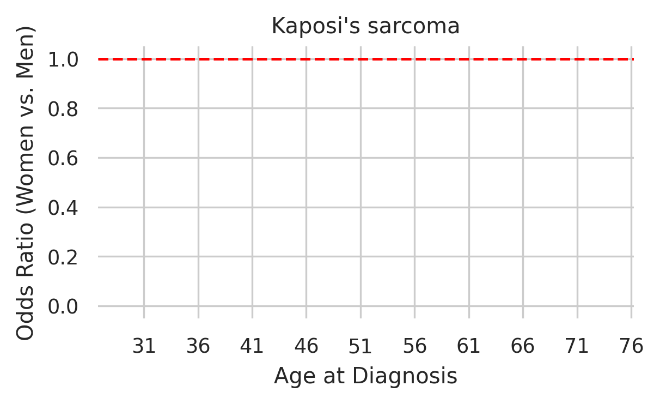**AQ**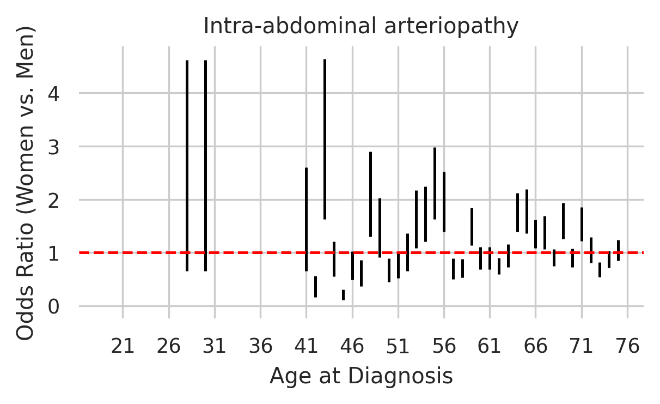 |
| **AR**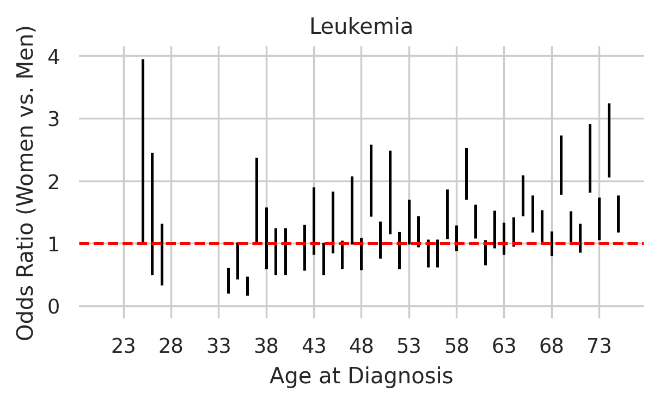 | **AS**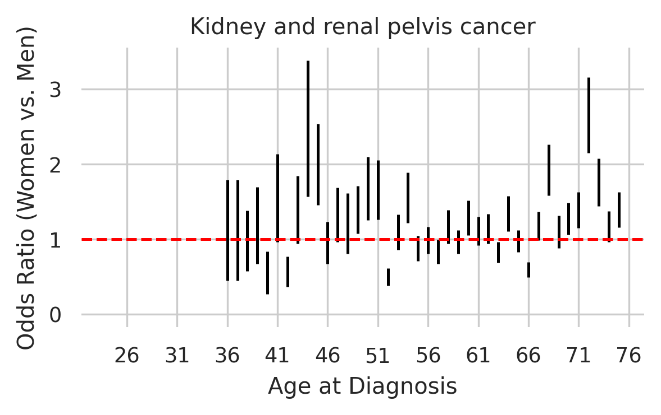 |
| **AT**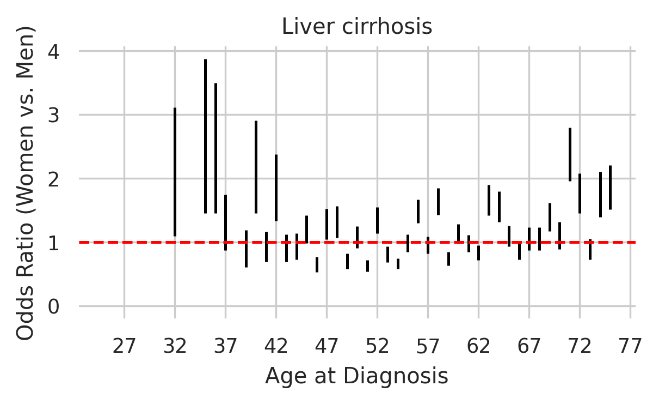 | **AU**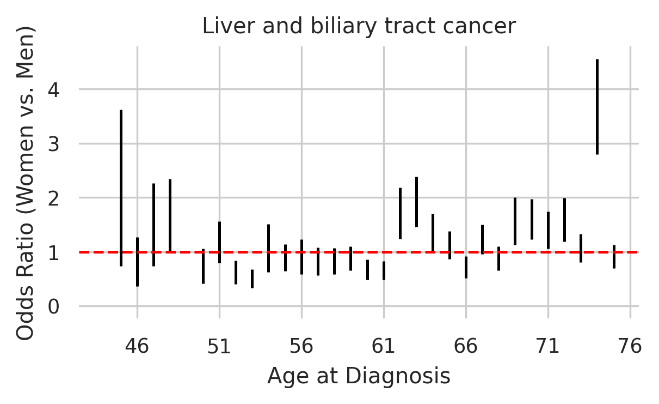 |
| **AV**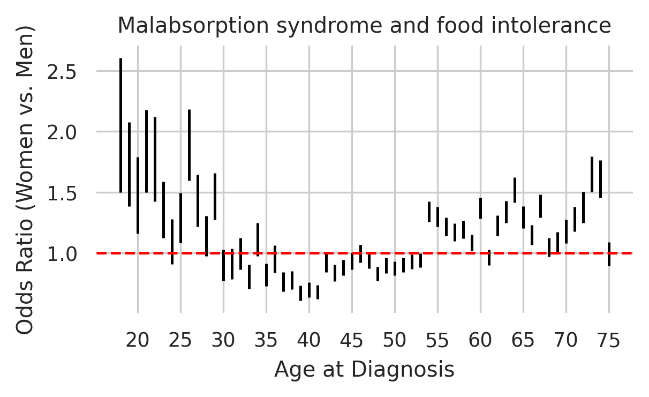 | **AW**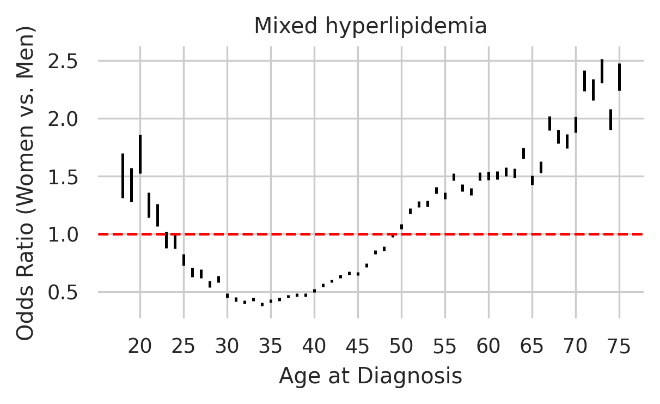 |
| **AX**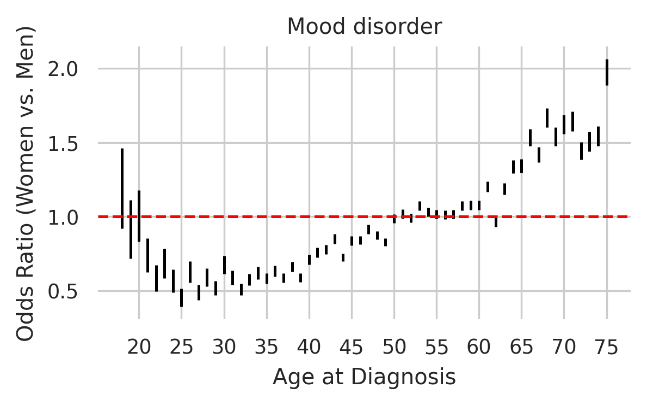 | **AY**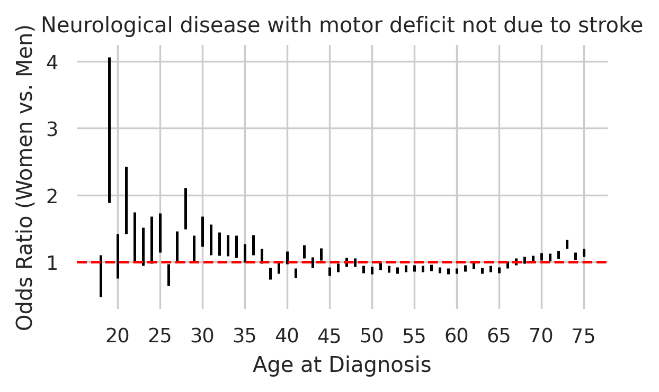 |
| **AZ**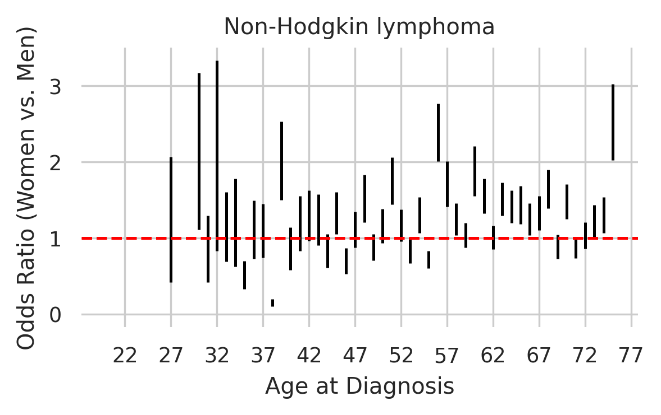 | **BA**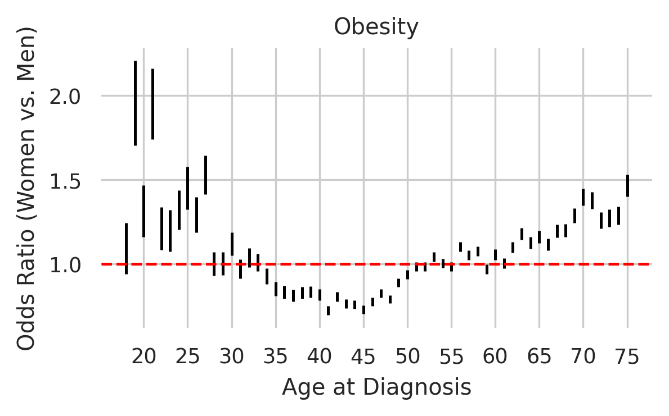 |
| **BB**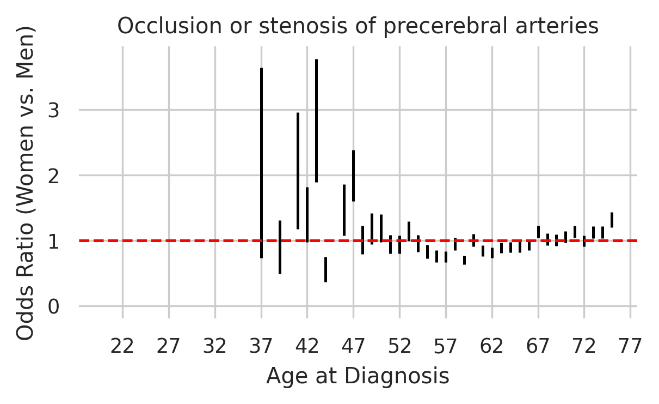 | **BC**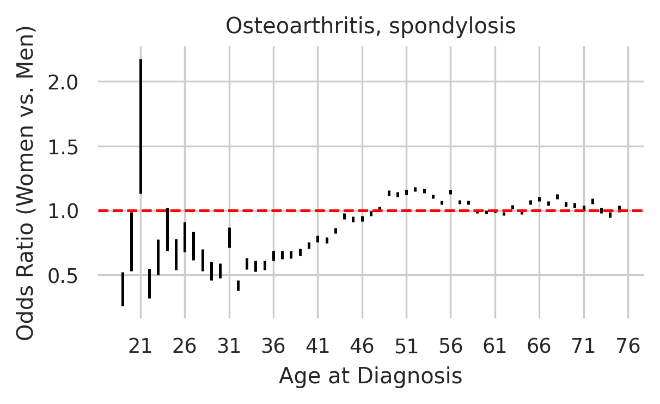 |
| **BD**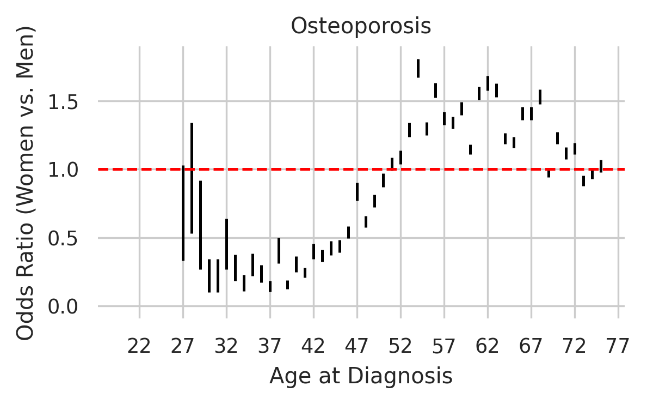 | **BE**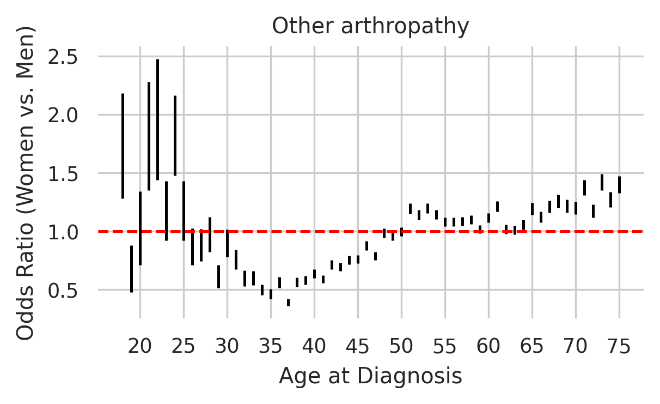 |
| **BF**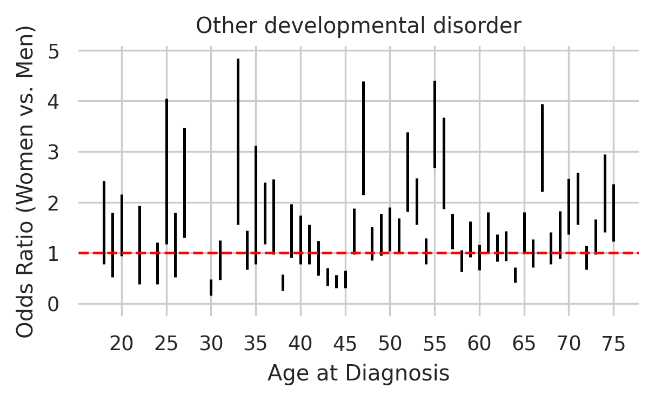 | **BG**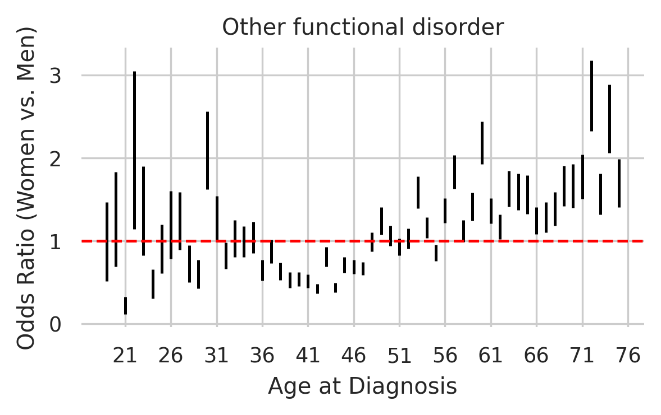 |
| **BH**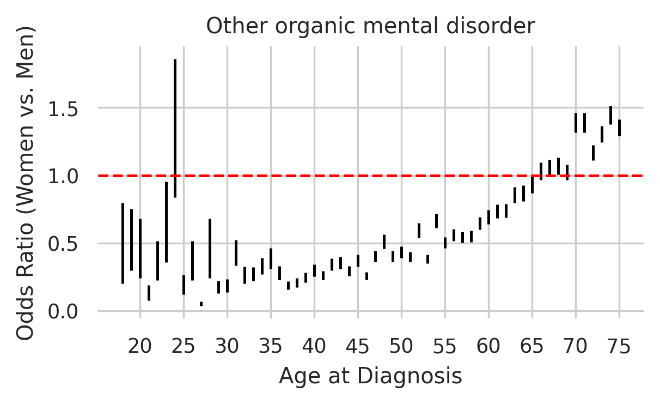 | **BI**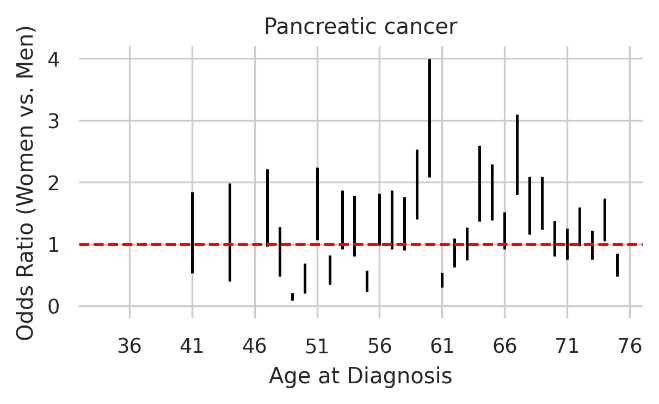 |
| **BJ**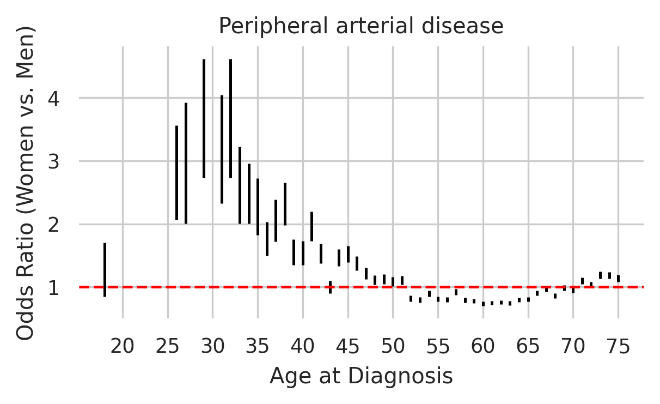 | **BK**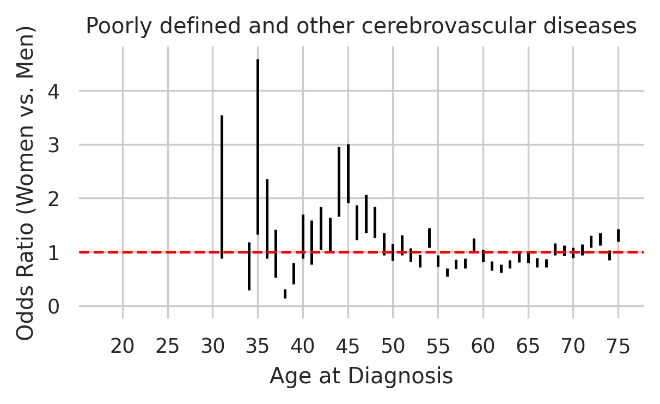 |
| **BL**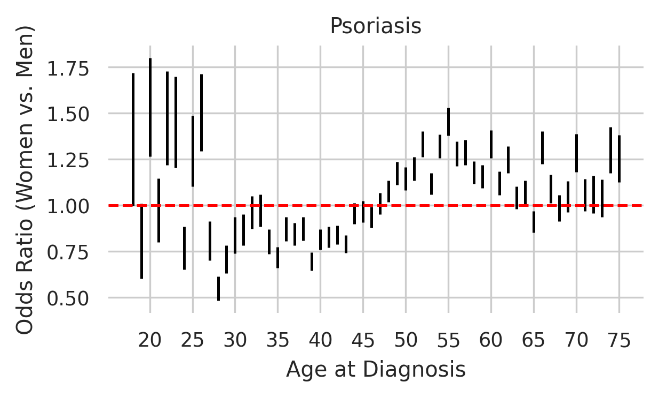 | **BM**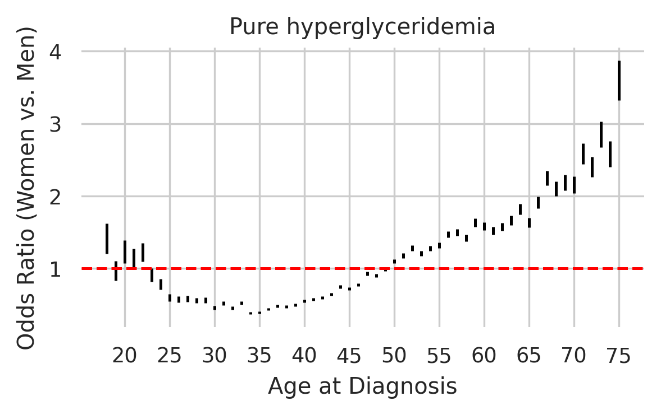 |
| **BN**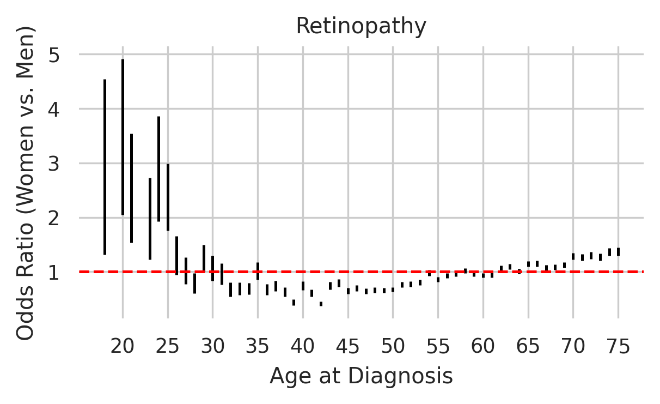 | **BO**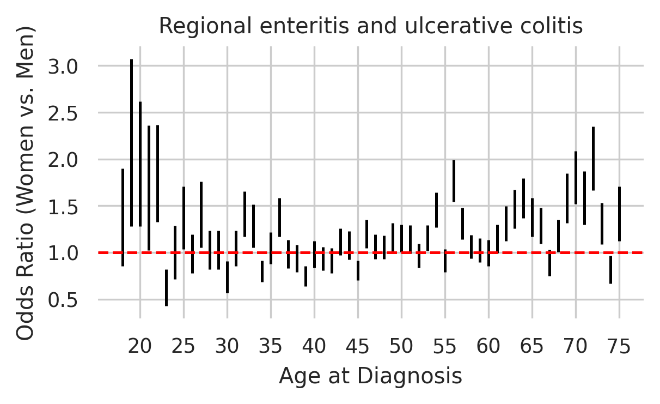 |
| **BP**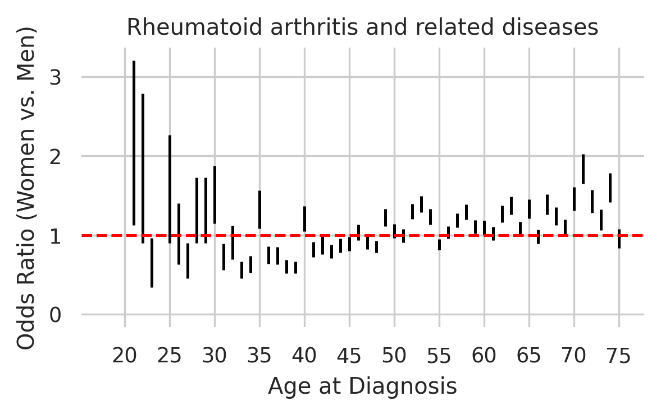 | **BQ**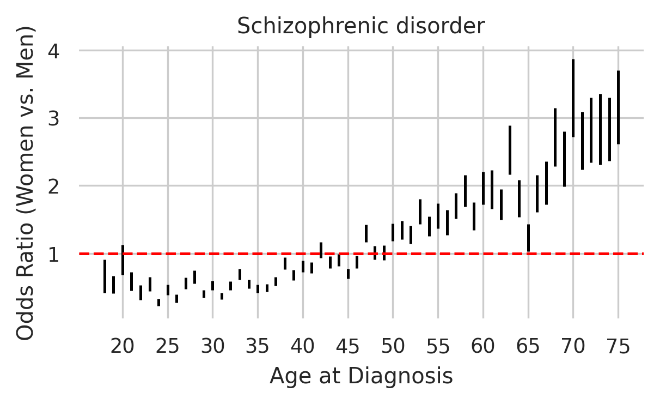 |
| **BR**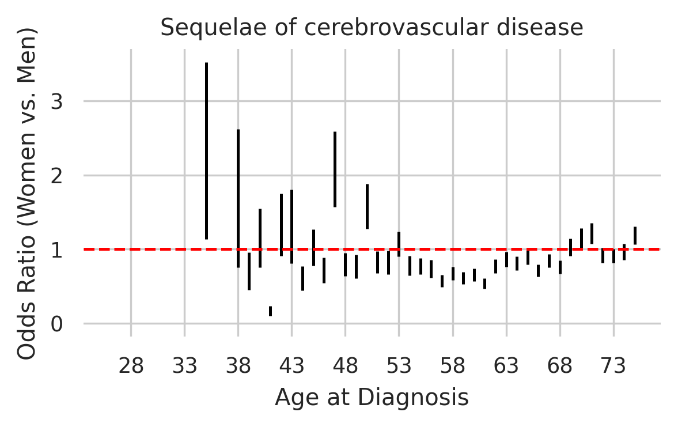 | **BS**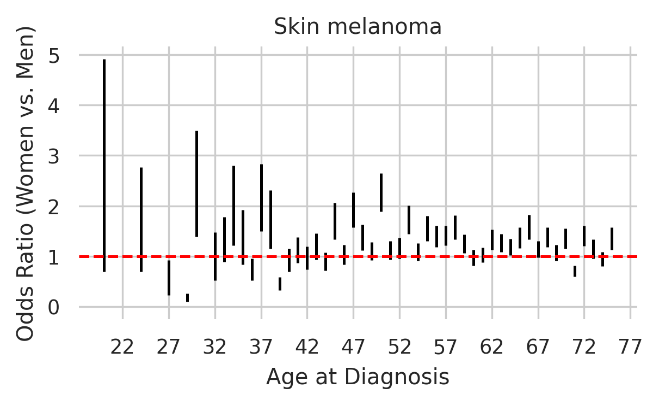 |
| **BT**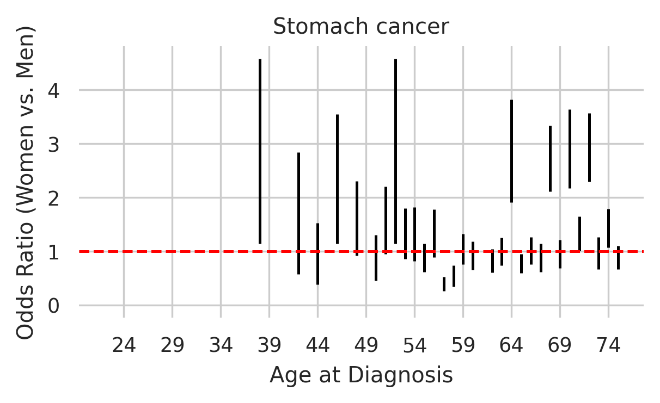 | **BU**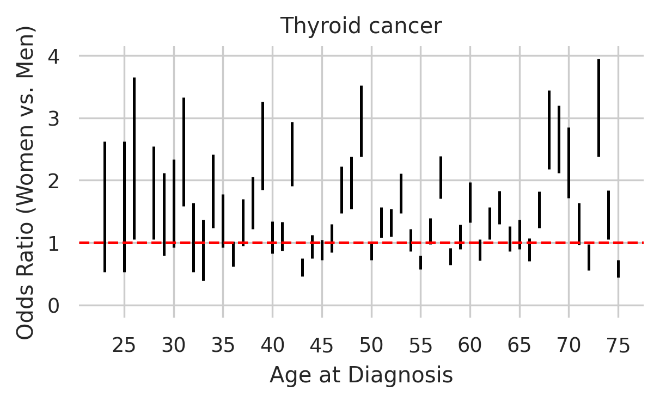 |
| **BV**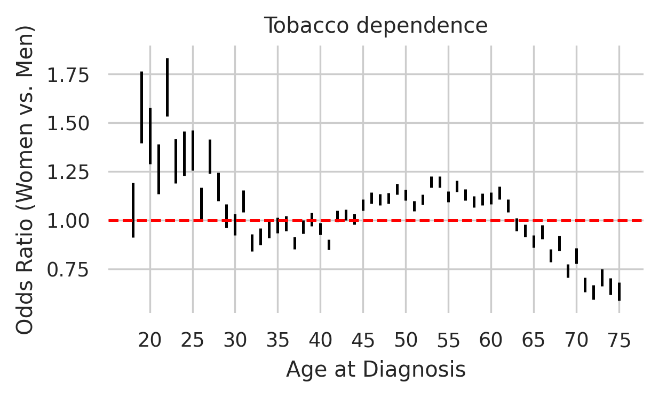 | **BW**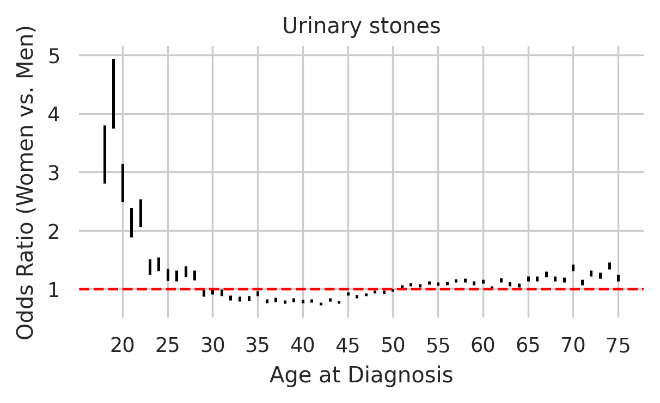 |
